# The Potential Public Health Impact of the mRNA-based Respiratory Syncytial Virus Vaccine, mRNA-1345, Under Extended Vaccination Campaigns Among Older Adults in the United Kingdom: A Modelling Study

**DOI:** 10.1101/2025.08.29.25334673

**Authors:** Mariia Dronova, Anna Tytula, Zuzanna Janusz, Parinaz Ghaswalla, Stuart Carroll, Orsolya Balogh, Keya Joshi

## Abstract

**Background/Objectives:** Respiratory syncytial virus (RSV) is a leading cause of severe respiratory disease in older adults. Despite growing recognition of RSV as a public health concern, vaccination options remain limited. This study assessed the potential long-term public health impact of extended mRNA-1345 RSV vaccination campaigns.

**Methods:** A dynamic transmission model, stratified by age, was developed to evaluate epidemiological and clinical impact of RSV vaccination in the UK over a 20-year time horizon. Eight vaccination strategies were assessed: two reflecting the JCVI recommendation for the 2024-2025 season and its recent extension, and six extended strategies considering broader eligible age groups, higher coverage, and/or revaccination every 2 or 3 years. Two exploratory analyses and extensive model validation versus reported data were also conducted.

**Results:** Strategies combining broader age eligibility (≥60 years), higher coverage (80%), and 2-year revaccination achieved the greatest impact, preventing 310,000 hospitalisations over 20 years in the total UK population. Exploratory analyses showed that the expected public health impact might exceed the estimates presented in this analysis, if an alternative vaccine efficacy profile or the projected demographic shift would be confirmed.

**Conclusions:** Extended RSV vaccination strategies including broader age eligibility and routine revaccination could offer substantial public health benefits in the UK. Targeting adults aged ≥60 years is expected to be particularly efficient in achieving a sustainable reduction in RSV burden. These findings could provide valuable support for national policy discussions on optimising RSV vaccination strategies in older adults, particularly regarding target age groups, revaccination schedules, and long-term programme planning.

## 1. Introduction

Respiratory syncytial virus (RSV) is one of the leading causes of acute respiratory tract infections in children and adults, with seasonal epidemics peaking in winter in temperate climates [1-3]. RSV usually causes a mild, self-limiting acute respiratory disease (ARD). However, infants, older adults, and adults with certain comorbidities are at higher risk of acute complications, particularly severe lower respiratory tract disease (LRTD), which may require hospitalisation and intensive care and can lead to death [1,3,4].

RSV affects an estimated 64 million people and results in up to 160,000 deaths per year worldwide [5]. Additionally, reinfections are common throughout life, including in old age [6,7]. In the United Kingdom (UK), cumulative incidence of RSV hospitalisations is reported at 43 and 65 cases per 100,000 persons over 65 years and over 75 years of age, respectively [1]. It is estimated that each winter, RSV causes 1200 deaths in persons aged 45 to 74 years and 4000 deaths in those aged over 75 years [1,8]. However, the true burden of RSV in older adults in the UK is likely under-recognised [9,10]. The annual cost to the National Health Service (NHS) for treating patients with RSV is substantial, and mostly attributable to individuals above 65 years. Thus, compared with younger adults, people aged 65 years and over are more likely to call NHS 111, more likely to consult their general practitioner, and are much more likely to be admitted to hospital. The direct cost of RSV cases in individuals ≥65 years is estimated at £93 million, which represents 67% of the total NHS expenditure for RSV management in all adults over age 18 years [11].

Diagnosis of RSV in older adults is challenging due to non-specific symptoms that overlap with other respiratory illnesses [11,12]. Additionally, hospitalisations and deaths in older adults with RSV are frequently attributed to underlying chronic conditions, such as chronic obstructive pulmonary disease (COPD) or heart failure, rather than RSV itself [13]. Timely identification of RSV infections could be also hindered by less frequent routine diagnostic testing for RSV in hospitalised older adults, when compared with paediatric patients, and reduced sensitivity of standard assays in this age group [6,12,14]. As a result, many cases remain undetected, leading to underreporting of RSV-related morbidity and mortality.

In response to growing evidence on the burden of RSV in the elderly, the Joint Committee on Vaccination and Immunisation (JCVI) recommended implementing an RSV vaccination programme for individuals aged 75-80 years, as a one-off campaign starting in September 2024, followed by routine immunisation of those turning 75 each year [1]. Recently, JCVI advised to extend the RSV vaccination campaign to adults aged 80 years and older, as well as to all residents in a care home for older adults [15]. This is aligned with the UK government’s wider “health mission”, which has placed a significant emphasis on prevention and vaccination [16]. It was also expected that such a programme could play an important role in alleviating pressure on the NHS, which routinely faces increased demand in winter due to respiratory infections. Indeed, RSV was mentioned as a key concern in NHS England’s Winter and H2 Priorities for 2024, highlighting the need for preventive measures to reduce hospitalisations and support healthcare capacity [17].

Treatment options for RSV are limited to supportive care, as no specific antiviral therapies are routinely recommended. Therefore, research and policy development are focused on preventive strategies, with vaccination playing a central role in reducing the burden of RSV for individuals and society. Recently, two new glycoprotein prefusion (PreF) vaccines and one ribonucleic acid (mRNA)-based PreF vaccine targeting RSV-LRTD were authorised by Medicines and Healthcare products Regulatory Agency (MHRA): Arexvy (RSVPreF3, GSK) [18,19], Abrysvo (RSVpreF, Pfizer), and mRESVIA (mRNA-1345, Moderna, Inc.) [20-22]. However, only the Pfizer vaccine (Abrysvo) was used during the first year of the UK immunisation campaign [23,24]. As evidence continues to emerge, the JCVI is now considering extensions to the programme, including the potential use of other vaccines, and broader eligibility criteria, including individuals at higher risk of severe illness and those younger than 75 years [24]. The safety and efficacy of mRNA-1345 in adults aged ≥60 years were confirmed in the pivotal Phase 2/3 ConquerRSV, a case-driven, randomised, double-blind, placebo-controlled, multi-continent study (NCT05127434) [25-27].

In light of recent policy developments and the availability of a new mRNA-based vaccine, there was a need to explore the comparative benefits of broad immunisation strategies targeting older adults. This study provides an assessment of the public health impact of extended vaccination campaigns with mRNA-1345 among adults aged ≥60 years in the UK, using a dynamic transmission model.

## 2. Materials and Methods

### 2.1 Study design

A dynamic transmission model (DTM) for RSV was developed, based on previously published model by Hodgson et al. [28,29]. The model captures key epidemiological features of RSV, including seasonality, age-dependent risk of RSV-associated ARD, LRTD, hospitalisations, and deaths; and a gradual build-up of natural immunity due to repeated exposures over a lifetime. The model was used to assess the potential public health impact of introducing an RSV vaccination campaign for older adults in the UK.

Population in the model was stratified into 18 age groups: 5-year age groups before 85 years, and a group 85 years and older. Demographic process allowed for simulation of population dynamics in the UK, reflecting changes in population size and population structure over time.

The analytical time horizon covered 20 RSV seasons, from September 2024 to August 2044. Prior to the start of the analytic time horizon, the model was run over a 30-year burn-in period (starting from 1994), to allow for stabilization of RSV transmission dynamics.

### 2.2 Model structure

Structure of the DTM developed for this analysis followed the established RSV modelling framework [9,28-32], comprising six main epidemiological compartments (M, S, E, I, A, and R), defined to follow the history of RSV infections and the acquired natural immunity over the lifetime. To capture the impact of vaccination, additional compartments (*V*) were introduced to allow tracking of vaccine-induced immunity and time post-vaccination. Model structure is presented in **Figure 1**.

**Figure 1.**
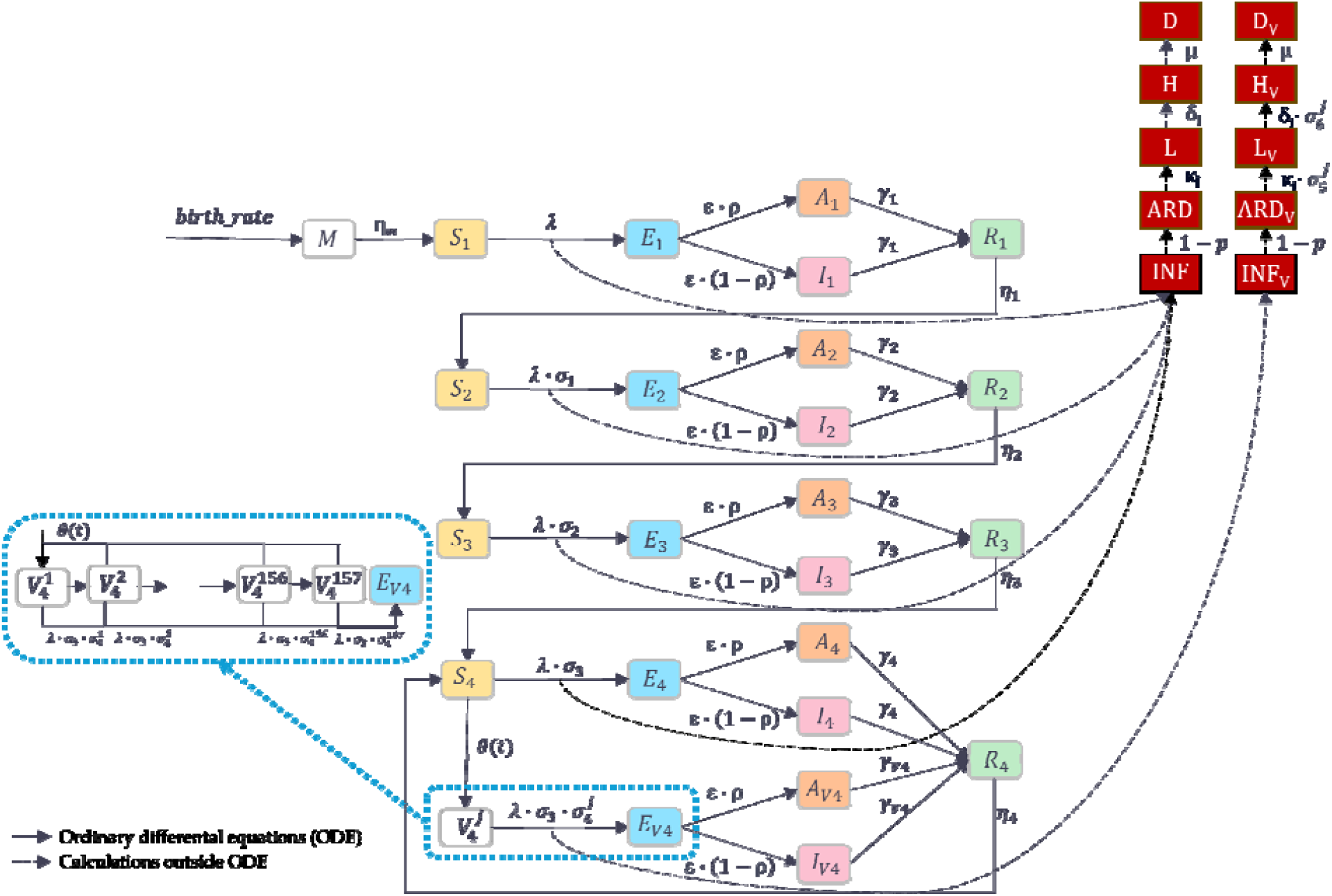
DTM structure. **Main compartments:** M – Maternal: maternal immunity from birth, S_i_ – Susceptible: susceptible to infection, E_i_ – Exposed: exposure to infection, A_i_ – Asymptomatic and infectious, I_i_ – Symptomatic and infectious (ARD), R_i_– Recovered: infection-induced immunity, – Vaccinated after 3+ infections, j-th week since vaccination, E_V4_ – Exposed: after vaccination, A_V4_ – Asymptomatic and infectious: after vaccination, I_V4_ – Symptomatic and infectious (ARD): after vaccination; for i, …, 4), j (1,…, 157) **Observational compartments:** INF/INF_V_ – Total number of RSV infections, ARD/ARD_V_ – Number of ARD cases, L/L_V_ – Number of LRTD cases, H/H_V_ – Number of hospitalised cases, D/D_V_ – Number of fatal cases **Main transmission parameters:** birth_rate – Birth rate, η_m_ – Waning rate of maternal immunity, λ – Force of infection, ε – Rate of becoming infectious, σ_i_ – Relative risk of re-infection, ρ – Proportion of infections that are asymptomatic, γ_i_ – Recovery rate from infection, η_i_ – Waning rate of post-infection immunity, κ_i_ – Proportion of LRTD among ARD, δ_i_ – Proportion of hospitalisations among LRTDs, μ – Proportion of deaths among hospitalized LRTD; for i (1,…, 4). **Vaccination parameters:** – Vaccination coverage, γ_V4_ – Recovery rate from infection in vaccinated, 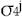 – Relative risk of RSV infection 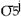 – Relative risk of LRTD given ARD 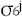 – Relative risk of hospitalisation given LRTD (in vaccinated vs not vaccinated n j-th

It was considered that individuals are born with the maternal protective immunity (*M*). Once this immunity wanes, an infant becomes susceptible to infection (*S*). The susceptible individual can acquire the infection and transition to an exposed but non-infectious state (*E*). Following a latent period, an infected individual becomes infectious, either with asymptomatic infection (*A*), or symptomatic infection, manifesting as ARD (*I*). Upon recovery from infection, an individual transitions to a recovery state (*R*), gaining a full protection against new infection due to natural immunity, which is maintained for a defined period of time. After this immunity wanes, an individual becomes susceptible to infection again (*S*). The model tracks individual across four exposure levels (i.e., first, second, third, and fourth or more infections), based on their RSV infection history.

Only individuals at the fourth exposure level were assumed eligible for vaccination, considering that the target population for immunisation included older adults. Following vaccination, an individual transitions from the susceptible state (*S*_4_) to the first compartment for vaccinated individuals 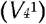. A vaccinated individual can experience a breakthrough infection, with the same disease pathway as described above, or transition to the next V compartment reflecting 1 week since vaccination, i.e., to 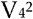 for the week 2 post-vaccination, then to 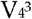 for the week 3 post-vaccination, etc., up to 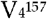 for the weeks 157+ post-vaccination. Vaccine-induced protection against RSV infection is applied for each V_4_ compartment, allowing to model waning of protection over time. Thus, the model allows to consider weekly changes of the level of protection over 3 years (156 weeks) since vaccination, and a stable protection for the following period (157+ weeks). The model also allows to consider protection against RSV-LRTD and hospitalisation, following breakthrough RSV infections.

In order to capture the clinical consequences of RSV infections, five observational compartments (*INF, ARD, L, H* and *D*) were defined to record the number of RSV infections, and associated cases of RSV-ARD, RSV-LRTD, RSV-related hospitalisation, and RSV-related death, respectively. Similarly, observational compartments *INF*_V_, *ARD*_V_, L_V_, *H*_V_, and *D*_V_ were defined for vaccinated individuals.

The main outcomes of the model were the total number of RSV infections, RSV-ARD cases, RSV-LRTD cases, hospitalisations and deaths as well as the total number of vaccine doses administered.

Further details on the modelling methodology are provided in Supplement 1.

### 2.3 Model calibration

The model was calibrated to the observed data using a two-step process, aiming to reflect the demographic and epidemiologic dynamics over the modelled timeframe, starting from 1994, which was the first year of the burn-in period.

The demographic calibration was performed to estimate the rate of population change, allowing to reproduce population growth and changes in age structure, as observed between 1994 [33] and 2023 [34]. This calibrated rate of population change was applied for the entire modelled time horizon and assumed to incorporate all-cause mortality and migration in the UK.

In addition, an exploratory scenario analysis was performed to account for potential faster change in population size and age distribution, considering the projection of population dynamics from the UK Office for National Statistics for 2030 [35].

The epidemiological calibration was performed to replicate the expected number of RSV-related hospitalisations by fitting three key transmission parameters: (1) the probability of transmission from an infectious to a susceptible individual, (2) the amplitude of the seasonal cosine function to capture variations in transmission intensity, and (3) the horizontal shift of the cosine function to align infection seasonality with the observed weekly patterns. Additionally, the proportion of RSV-LRTD among RSV-ARD was adjusted during the calibration process to improve the fit to age distribution of hospitalisation rate.

Further details on the model calibration are provided in **Supplement 1**.

### 2.4 Model inputs

Demographic process in the model was informed by the data on population size and birth rates reported by the UK Office for National Statistics [33-38]. The total number of physical and conversational contacts between different age groups by week was calculated using the data from the POLYMOD study [39] and the R package socialmixr [40].

Calibration of transmission parameters and dynamics of RSV incidence was informed by the hospitalisation rate per 100,000 individuals in the total population. Hospitalisation rates varied across age groups and by week of the year, following a characteristic seasonal pattern with a winter peak. Age-specific hospitalisation rates for adults aged ≥65 years were derived from results of a recent large-scale study published by Osei-Yeboah et al. [41], providing detailed estimates for this population. However, as this source did not report data for individuals aged 0–17 years and only provided aggregated estimates for 18 to 64 year-olds, hospitalisation rates for those aged 0–64 years were based on the UK national surveillance report [42], scaled up by an underreporting factor of 1.5 derived from McLaughlin et al. [43]. This adjustment was necessary due to the sentinel nature of the surveillance system in the UK, which likely underestimates the true burden of RSV [1,44]. It should be noted that the underreporting of RSV hospitalisations in elderly has been recognised in the UK as well as for other countries. For example, a recent modelling study for Germany considered much higher underreporting factors in a range of 8-14 [9]. Seasonal distribution of hospitalisation rate was informed by the national surveillance report data from week 17 of 2023 to week 16 of 2024 [42].

Severity of an incident RSV infection in the model was informed by four key parameters: (1) the proportion of ARD among all RSV infections [9,45], (2) the proportion of RSV-LRTD among RSV-ARD cases [calibrated], (3) the proportion of RSV hospitalisations among RSV-LRTD cases [43,46-48], and (4) the proportion of deaths among hospitalized cases [46,47,49]. All severity parameters were age-specific, in line with the model’s age stratification, where the data availability allowed to consider sufficient granularity. Duration of maternal immunity, latency period, infection and antibody protection for those previously infected as well as relative risk of infection and relative infectiousness were informed by Hodgson et al. [28], and complemented by the data from Pitzer et al. [50].

Further details on the model input values are provided in **Supplement 1**.

### 2.5 Vaccination parameters

Vaccine efficacy was informed by results of a Phase 2/3 ConquerRSV trial [25,27]. In the main analysis, vaccination with mRNA-1345 was considered to provide two types of protection: (1) protection against RSV infection, including asymptomatic and ARD, and (2) protection against RSV-LRTD. Protection against asymptomatic infection was assumed equal to protection observed against ARD. For each type of protection, non-linear waning was applied for 3 years post-vaccination, and was conservatively set to zero for the following period, in the absence of long-term data on vaccine-derived immunity [51].

In an exploratory analysis, an alternative assumption was tested, considering an additional type of protection against RSV-related hospitalisations, and linear waning for all three types of protection. Similarly to the main analysis approach, the 3-year duration of protection was applied.

Vaccine efficacy in the first week post-vaccination and mean efficacy values for the following years are presented in Table 1. A detailed description of the methodology used to derive vaccine efficacy estimates is provided in **Supplement 1**.

**Table 1.**
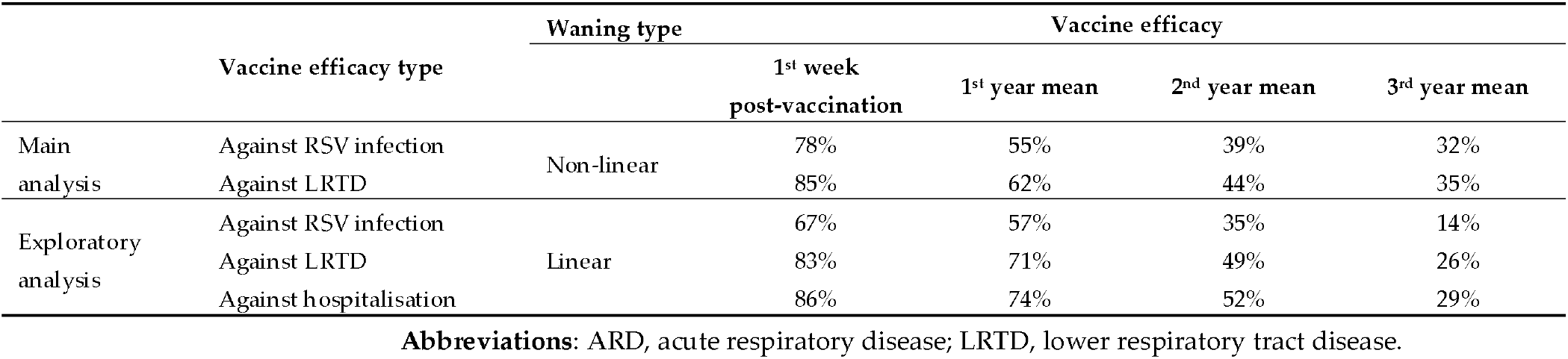
Estimated vaccine efficacy.

Eight vaccination strategies were explored, including Strategy 1, reflecting the JCVI recommendation for the 2024-2025 season [1]; Strategy 2, mirroring an updated JCVI recommendation, covering also individuals ≥ 80 years old [15]; and six extended strategies, considering broader eligible age group, higher coverage, and/or revaccination every 2 or 3 years. A summary of the evaluated vaccination strategies is provided in **Table 2**.

**Table 2.** Overview of the vaccination strategies included in the analysis.

| Strategy | Eligible age group <sup>1</sup> | Target coverage <sup>2</sup> | Revaccination |
| --- | --- | --- | --- |
| Strategy 1 | 75-80 years | 60% | No |
| Strategy 2 | $\geq 75$ years | 60% | No |
| Strategy 3 | 75-80 years | 60% | Every 2 years |
| Strategy 4 | $\geq 60$ years | 80% | Every 2 years |
| Strategy 5 | $\geq 60$ years | 60% | No |
| Strategy 6 | 75-80 years | 80% | No |
| Strategy 7 | 75-80 years | 60% | Every 3 years |
| Strategy 8 | $\geq 60$ years | 80% | Every 3 years |

<sup>1</sup> Routine vaccination for the first single-year cohort of the eligible age group was included for all strategies e.g., strategy 1 considered vaccination for individuals aged 75–80 years in the first year of the campaign and for individuals turning 75 years each next year.
<sup>2</sup> A target coverage rate of 60% was based on the observed uptake of RSV vaccine for the first season of vaccine introduction, as of 31 March 2025 [52]. A target coverage of 80% was informed by influenza vaccine uptake among individuals $\geq 65$ years old during the 2024–2025 season [53]. It was assumed that vaccine uptake occurs from week 36 to week 52 of the year (September to December), with the weekly uptake pattern informed by the data for influenza vaccine for the 2024–2025 season [54]. Further details on the input values are provided in **Supplement 1**.

### 2.6 Model validation

Following modelling best practices [55], the DTM underwent a comprehensive validation process. Model predictions for key outcomes (per-capita probability of transmission, age-specific ARD and hospitalisation rate, the expected impact of vaccination) were compared with published findings from other models [28,46,56-59]; and relevant literature [11,12,28,29,42,60].

## 3. Results

### 3.1 Model fit

The model calibration achieved a fairly good fit to the population structure in the UK and the observed dynamics of RSV-related hospitalisations. For the starting year of the analysis (2024), the difference between the reported and simulated size of the total population was 1%, and up to 4% across age groups. Similarly, simulated hospitalisation rates were aligned with the reported epidemiological data, capturing the overall incidence, age distribution, and seasonality of the disease. The difference between the expected and simulated number of hospitalizations was up to 5% for the total population and across age groups (considering the number of hospitalizations over a calibration timeframe).

Further details on the calibration outputs are provided in **Supplement 2**.

### 3.2 Validation

Model outputs were assessed against published estimates to evaluate the plausibility of simulated disease dynamics and impact of vaccination.

The probability of transmission per contact in the presented model was estimated at approximately 8%, which was reasonably close to the value of 10% reported by Hodgson et al. [28], with variation likely reflecting differences in model parameterisation.

Predicted age-specific incidence rates of ARD were in line with estimates reported in the literature across most age groups (**Supplement 2, Table S15**). Published values vary widely due to heterogeneity in study design, case definitions, and quality of surveillance. For example, the DTM-predicted that average annual ARD incidence in adults aged ≥18 years was 7,321 per 100,000, falling within the range of 900–13,100 reported by Wilkinson et al. [12], and in line with the RAND estimate of 6,481 per 100,000 [11]. Similarly, for older adults, model predictions were well aligned with these studies [11,12].

Predicted age-specific hospitalisation rates were within the reported ranges (**Supplement 2, Table S16**). For individuals aged ≥65 years, the simulated hospitalisation rate of 181 per 100,000 persons was higher than that reported by United Kingdom Health Security Agency (UKHSA) (59 per 100,000 persons) [42], likely reflecting differences in data collection methodologies and not accounting for underreporting due to suboptimal diagnostic tests. The hospitalization rate was also higher than those reported by Howa et al. (91 per 100,000 persons) [56]. However, the simulated rate aligned well with the estimate of about 160 per 100,000 persons from Sharp et al. [59], Hodgson et al. [29], and RAND [11], while remaining lower than the upper bound of 193 per 100,000 from Fleming et al. [46], 225 per 100,000 from Johannesen et al. [57], and 276 per 100,000 from Zhang et al. [60].

Overall, results of the model validation for no vaccination arm indicate that the model reproduces key patterns of RSV epidemiology well and provides a reasonable basis for assessing the public health impact of vaccination strategies.

Ultimately, as the first evidence on the real-world effect of RSV vaccination from the 2024-2025 pilot campaign became available [58], model outputs were compared with the results of this study. A dedicated validation scenario was conducted to align the modelled vaccine coverage with that captured in the study [58], considering the observed RSV vaccine uptake in 2024-2025 season, up to the first week of 2025 [52]. Under this validation scenario, the model predicted a 26% reduction in RSV-related hospitalisations, closely matching the 30% reduction (95% CI: 18–40%) observed in the real-world study [58], despite the differences in the vaccine brand considered. This alignment with observed outcomes under real-world conditions supports the plausibility of the model’s projections and its use in evaluating long-term vaccination strategies.

### 3.3 RSV vaccination strategies: impact in the targeted population

Vaccination against RSV was shown to provide considerable benefits by significantly reducing the burden of disease over 20 years in the target population of older adults in the UK.

Results of the analysis are summarized in **Table 3** and **Table S17** for the main clinical outcomes: RSV-ARD, RSV-LRTD, RSV-related hospitalisations, and deaths. While the numeric estimates are presented across all outcomes, the further description focuses on hospitalisations as a representative measure of RSV burden.

**Table 3.** Number of cases, number of cases avoided, and percentage of cases avoided in the target population, mRNA-1345 vs. no vaccination over 20 years

| Strategy | No vaccination |  |  | Strategy 1 | Strategy 2 | Strategy 3 | Strategy 4 |
| --- | --- | --- | --- | --- | --- | --- | --- |
| Age group | 75-80 y | ≥ 75 y | ≥ 60 y | 75-80 y | ≥ 75 y | 75-80 y | ≥ 60 y |
| Coverage | - | - | - | 60% | 60% | 60% | 80% |
| Revaccination | - | - | - | No | No | Every 2 y | Every 2 y |
| **Number of cases** |  |  |  |  |  |  |  |
| RSV ARD | 4,195,188 | 7,128,679 | 20,053,473 | 3,621,374 | 6,361,525 | 2,845,656 | 10,852,108 |
| RSV LRTD | 997,987 | 2,226,664 | 3,725,497 | 845,624 | 1,994,585 | 640,674 | 1,784,194 |
| RSV hospitalisation | 186,227 | 415,502 | 552,758 | 157,796 | 372,195 | 119,552 | 259,137 |
| RSV death | 123,356 | 275,226 | 318,698 | 104,523 | 246,540 | 79,190 | 147,222 |
| **Number of cases avoided** |  |  |  |  |  |  |  |
| RSV ARD | - | - | - | 573,814 | 767,154 | 1,349,532 | 9,201,364 |
| RSV LRTD | - | - | - | 152,363 | 232,079 | 357,313 | 1,941,303 |
| RSV hospitalisation | - | - | - | 28,431 | 43,307 | 66,676 | 293,622 |
| RSV death | - | - | - | 18,833 | 28,686 | 44,165 | 171,475 |
| **Percentage of cases avoided** |  |  |  |  |  |  |  |
| RSV ARD | - | - | - | 14% | 11% | 32% | 46% |
| RSV LRTD | - | - | - | 15% | 10% | 36% | 52% |
| RSV hospitalisation | - | - | - | 15% | 10% | 36% | 53% |
| RSV death | - | - | - | 15% | 10% | 36% | 54% |

**Abbreviations:** ARD, acute respiratory disease; LRTD, lower respiratory tract disease; y, years; RSV, respiratory syncytial virus; y, years.
**Note:** Percentages of cases avoided are calculated using the total number of cases without vaccination as a denominator, for each respective target age group. Due to variation in population size, percentages are not directly comparable between strategies with different eligibility criteria.

Across all explored strategies, the number of hospitalisations avoided varied from 28,000 to 294,000 (7-53% cases avoided) in the target population, depending on the strategy design.

A one-time campaign targeting adults aged 75–80 years, reflecting the pilot UK programme for 2024-2025 (Strategy 1), was projected to provide a limited reduction of RSV burden over the long term, due to the narrow age range considered and absence of revaccination. This strategy was estimated to prevent more than 28,000 hospitalisations in the target population.

Other strategies, exploring the impact of the vaccination programme extensions (broader eligibility, higher coverage, and/or revaccination), are expected to further reduce the RSV burden.

Strategy 2, which reflects the updated 2025 JCVI guidance on inclusion of individuals >80 years in the campaign, resulted in a slightly higher impact than Strategy 1, preventing more than 43,000 hospitalisations in the target age group, over a 20-year period.

A distinctly larger reduction in RSV burden was observed for strategies with revaccination, in contrast to one-time campaigns described above. Thus, Strategy 3, considering revaccination every 2 years, even with 60% coverage and the narrow eligibility criteria (age group of 75-80 year-olds), was associated with more than 66,000 hospitalisations avoided.

Strategy 4, which combined broad age eligibility (≥60 years), reasonably high vaccine uptake (80%), and repeated vaccination every 2 years, was shown to be the most beneficial, as might be expected (**Table 3**). This strategy prevented more than 293,000 hospitalisations in the target population. The magnitude of the projected impact highlights the advantage of maintaining protection through repeated vaccination and broader eligibility criteria.

Results for the Strategies 5–8 are shown in **Table S17**.

### 3.4 RSV vaccination strategies: impact in the total population

In addition to reducing disease burden in the target population, RSV vaccination in older adults was also expected to provide tangible clinical benefits at the total population level, with 36,000 to ∼310,000 (3-28%) hospitalisations avoided, depending on the strategy (**Table 4 and Table S18**). The projected effect in the total population was mainly attributed to the direct protection in the vaccinated age groups, with the most substantial impact achieved with strategies including broader age eligibility, higher coverage, and repeated vaccination. Particularly, Strategy 4 was projected to prevent ∼310,000 hospitalisations in the total population, over 20-year time horizon. Results are presented in **Figure 2**.

**Table 4.** Number of cases, number of cases avoided, and percentage of cases avoided in the total population, mRNA-1345 vs no vaccination, over 20 years

| Strategy | No vaccination | Strategy 1 | Strategy 2 | Strategy 3 | Strategy 4 |
| --- | --- | --- | --- | --- | --- |
| Age group | Total population | 75-80 y | ≥ 75 y | 75-80 y | ≥ 60 y |
| Coverage | - | 60% | 60% | 60% | 80% |
| Revaccination | - | No | No | Every 2 y | Every 2 y |
| **Number of cases** |  |  |  |  |  |
| RSV ARD | 153,877,864 | 152,862,383 | 152,690,490 | 150,480,043 | 139,389,096 |
| RSV LRTD | 12,025,486 | 11,815,059 | 11,764,172 | 11,213,520 | 9,795,240 |
| RSV hospitalisation | 1,090,248 | 1,053,760 | 1,044,869 | 948,711 | 780,471 |
| RSV death | 336,077 | 312,687 | 306,969 | 245,125 | 163,511 |
| **Number of cases avoided** |  |  |  |  |  |
| RSV ARD | - | 1,015,481 | 1,187,373 | 3,397,821 | 14,488,767 |
| RSV LRTD | - | 210,427 | 261,313 | 811,965 | 2,230,246 |
| RSV hospitalisation | - | 36,488 | 45,378 | 141,537 | 309,777 |
| RSV death | - | 23,390 | 29,109 | 90,952 | 172,566 |
| **Percentage of cases avoided** |  |  |  |  |  |
| RSV ARD | - | 1% | 1% | 2% | 9% |
| RSV LRTD | - | 2% | 2% | 7% | 19% |
| RSV hospitalisation | - | 3% | 4% | 13% | 28% |
| RSV death | - | 7% | 9% | 27% | 51% |

**Abbreviations:** ARD, acute respiratory disease; LRTD, lower respiratory tract disease; RSV, respiratory syncytial virus; y, years.

**Figure 2.**
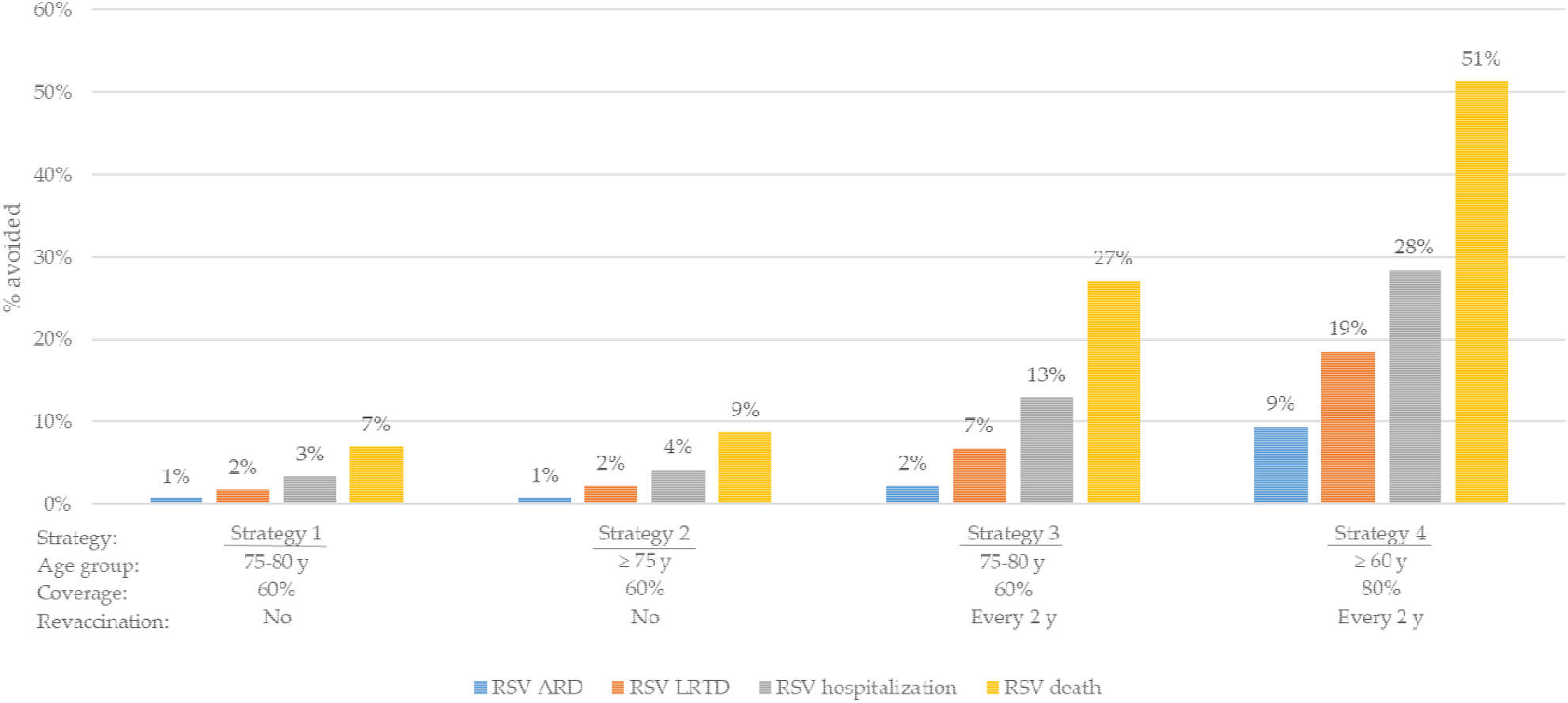
Percentage of cases avoided in total population, mRNA-1345 vs no vaccination (over 20 years). **Abbreviations:** ARD, acute respiratory disease; LRTD, lower respiratory tract disease; RSV, Respiratory syncytial virus; y, years.

Beyond the direct effect in vaccinated individuals, decrease in RSV-related morbidity and mortality were also expected in non-target age groups (**Figure 3Error! Reference source not found**.), due to indirect protection driven by the modelled reduction in infection risk among vaccinated individuals. Notably, the largest indirect effects were observed in adjacent age cohorts — particularly those just below the vaccination threshold (e.g., for 55–59 year-olds in Strategy 4, targeting adults ≥60 years old). This reflects the realistic contact patterns between adjacent age groups, and the corresponding decrease in exposure to infection among older adults who may be at increased risk of severe RSV disease, but are not yet eligible for immunisation.

**Figure 3.**
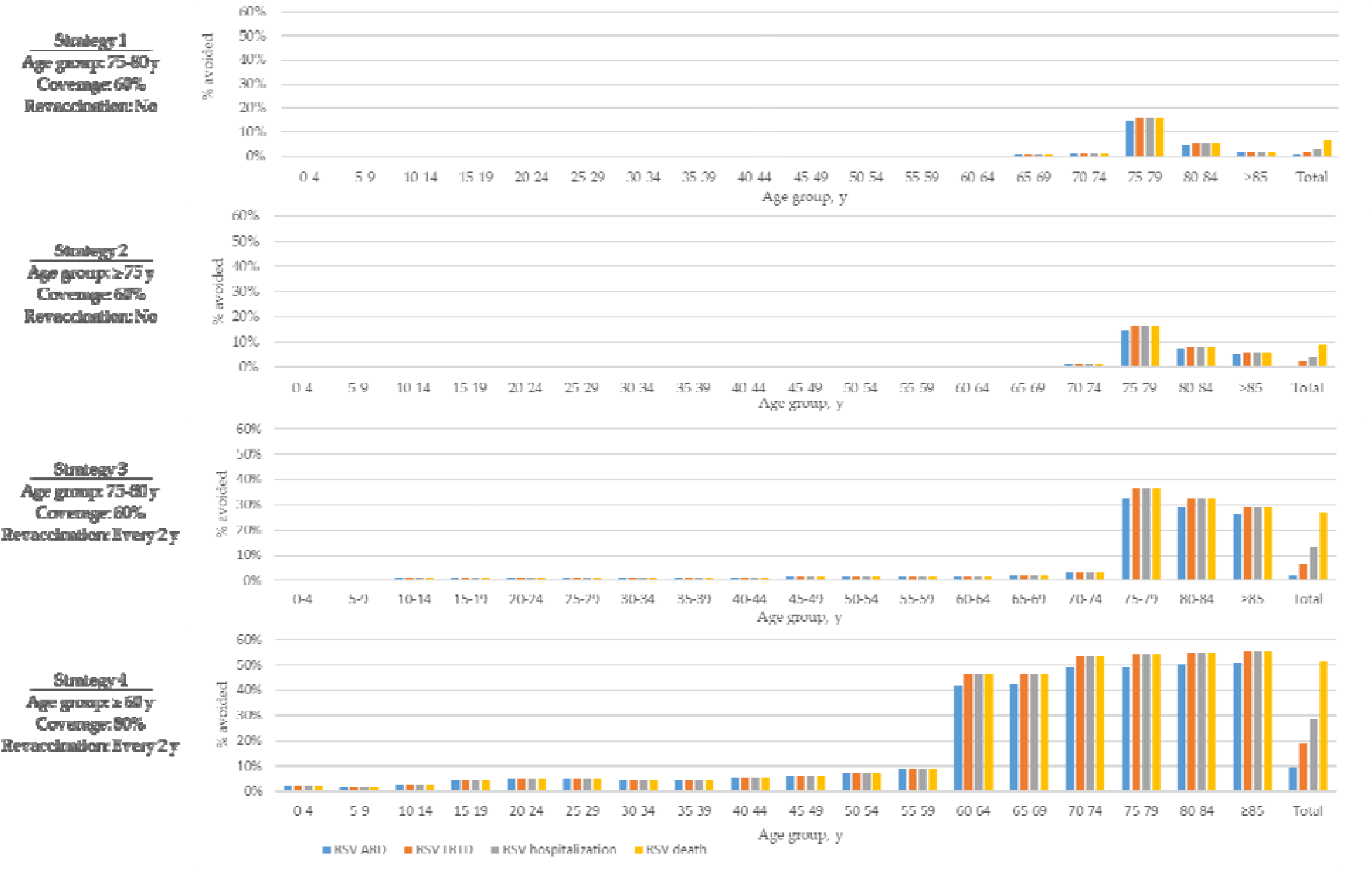
Percentage of cases avoided in total population by age group, mRNA-1345 vs no vaccination (over 20 years). **Abbreviations:** ARD, acute respiratory disease; LRTD, lower respiratory tract disease; RSV, respiratory syncytial virus; y, years.

Differences in population-level impact across strategies reflected the modelled vaccination strategy design, particularly the duration of vaccine protection and inclusion of revaccination. The temporal trends over the 20-year horizon (**Figure 4**

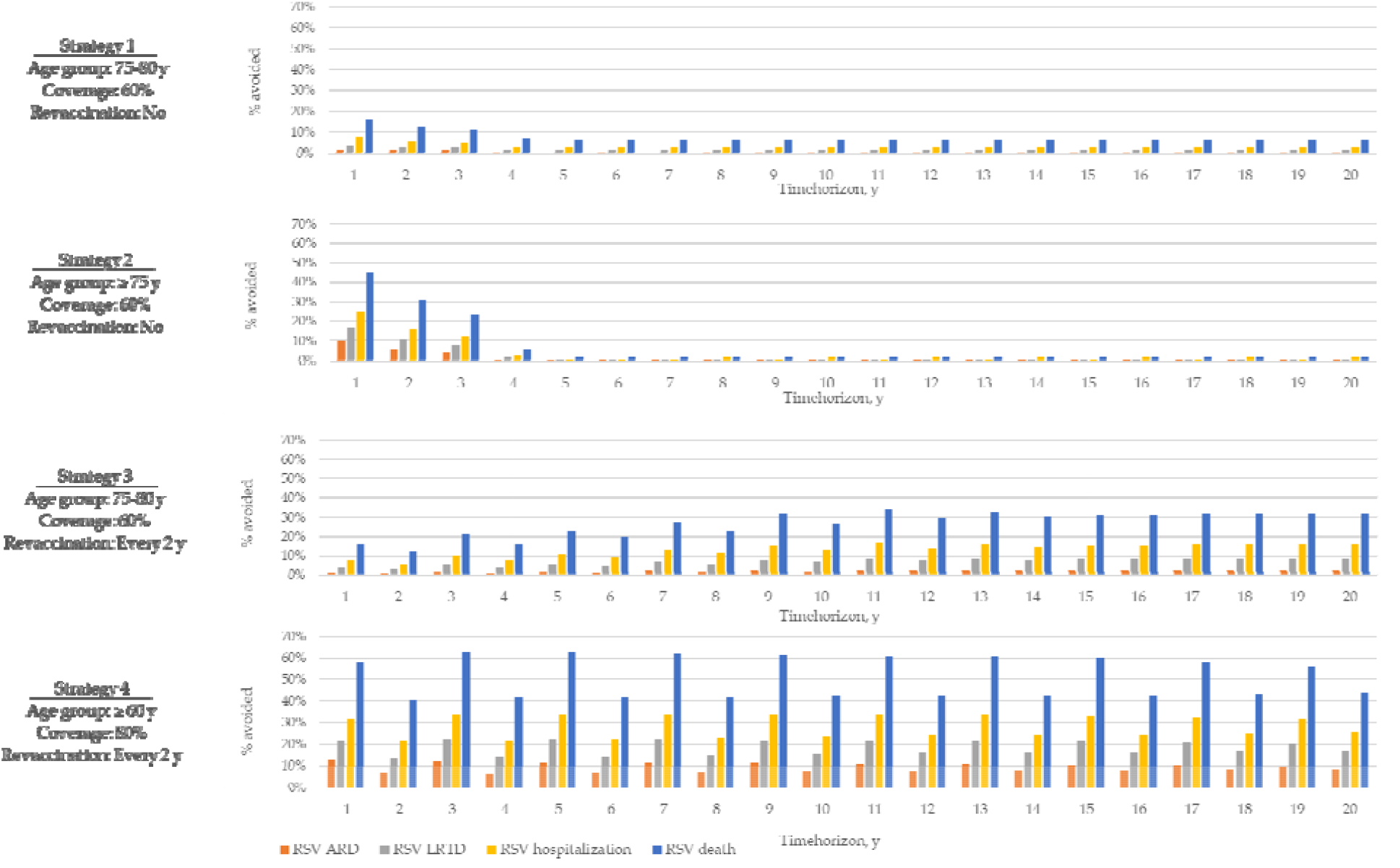

**Figure 4**) show distinct differences between one-time and continuing vaccination approaches. In Strategies 1 and 2 based on one-time vaccination, the clinical benefits were concentrated within the first 3 years after the start of the immunisation programme, with a low disease impact over the following period. The early impact was driven by the initial vaccination of the full eligible cohort in year 1 and the assumed duration of vaccine-induced protection (up to 3 years). After this period, the effect declined substantially, as protection in the initially vaccinated cohort waned, and the strategy transitioned to a maintenance phase, in which vaccination was provided only to individuals within a single-year age cohort who became newly eligible. As a result, the disease impact plateaued at a low level, leading to limited cumulative gains over the long term.

**Figure 4.**
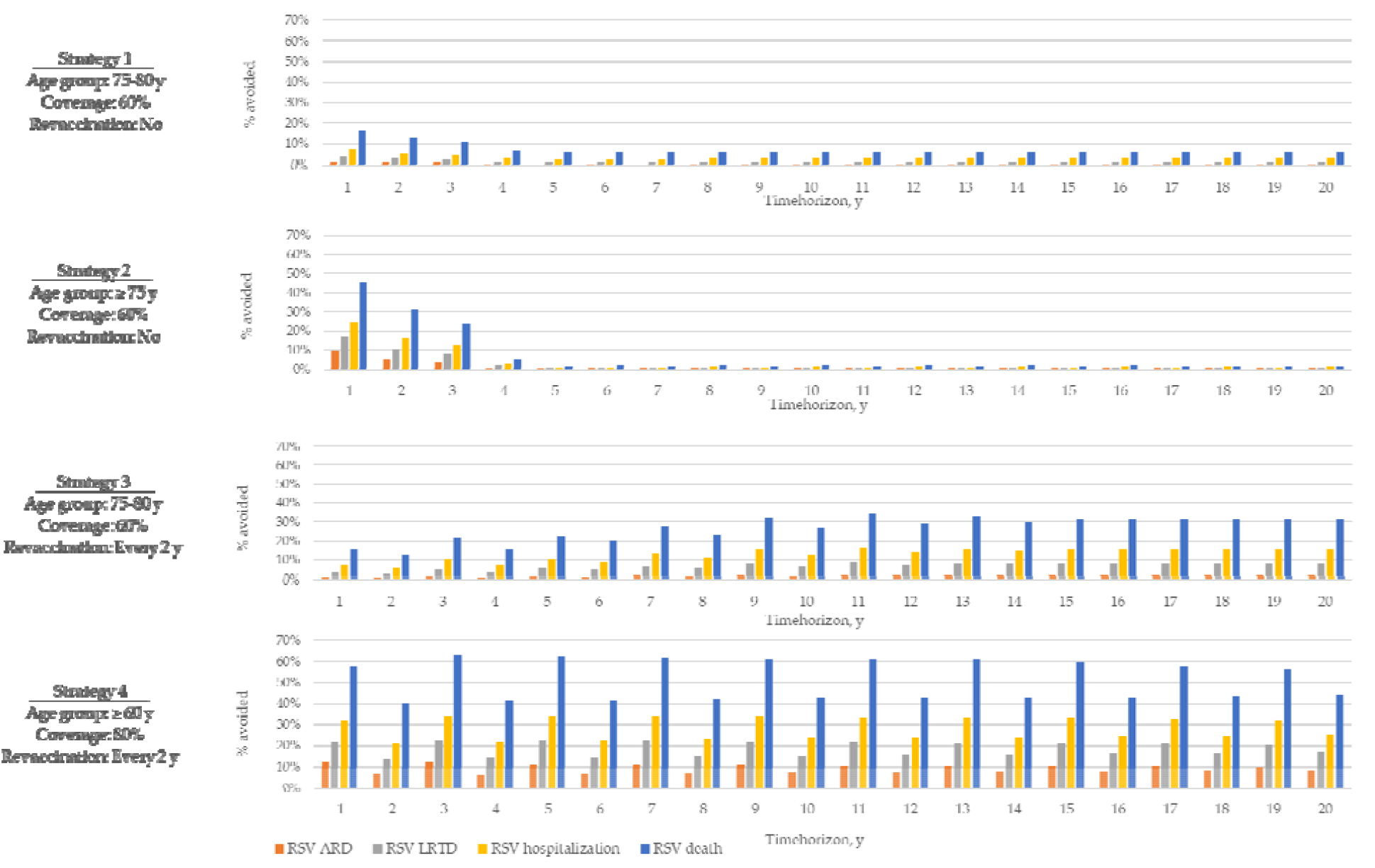
Percentage of cases avoided in total population over time, mRNA-1345 vs no vaccination. **Abbreviations:** ARD, acute respiratory disease; LRTD, lower respiratory tract disease; RSV, respiratory syncytial virus; y, years.

In contrast, Strategies 3 and 4 which included repeated vaccination at regular intervals demonstrated a distinctly different temporal pattern, with clinical benefits more evenly distributed across the 20-year model horizon (**Figure 4**). These strategies allowed maintained protection against RSV over time, with the most persistent reductions observed under Strategy 4. Among the repeated-dose strategies, vaccination every second year resulted in the greatest and most stable reductions across all outcomes. This approach allowed the level of protection to be restored before it fully waned, providing a sustained benefit for the target and total population level.

Results for the Strategies 5–8 are shown in **Table S18** and **Figures S11–S13**

### 3.5 Exploratory analysis

Simulated projections of long-term impact across strategies are shaped by several assumptions, particularly those related to vaccine protection and demographic trends. In the present analysis, two alternative scenarios were examined to test the effect of (1) inclusion of protection against hospitalization and a linear waning profile, and (2) accelerated ageing and population size increase. An alternative scenario on vaccine efficacy aimed to test the impact of differences in short-term protection dynamics, in absence of long-term evidence on duration of protection. A scenario on demographic shift was designed to reflect a projected increase in the size of a target population, which may be associated with a growing burden of RSV over time.

Although the future epidemiology of RSV remains uncertain, current estimates of its burden in older adults also carry considerable uncertainty due to limitations in reporting. While the absolute number of RSV-related clinical events may vary under different epidemiological assumptions, the relative performance of the evaluated vaccination strategies is expected to hold, particularly the comparative benefits of a broader age eligibility and repeated dosing. Therefore, the dedicated exploratory analysis on epidemiological parameters was not conducted, and further considerations regarding this area of uncertainty and its implications are described further in sections 3.5 and 4.

Exploratory analyses were conducted for Strategies 4 and 8 only. The observed trends in disease impact were similar for these two strategies, therefore the results for Strategy 4 are presented below, with corresponding outputs for Strategy 8 provided in **Supplement 2, Table S20**.

The disease impact of Strategy 4 under exploratory scenarios was comparable to that estimated for the main analysis (Table 5), although the results illustrate potential shifts in the expected outcomes of vaccination programme that could emerge as further evidence becomes available.

**Table 5.**
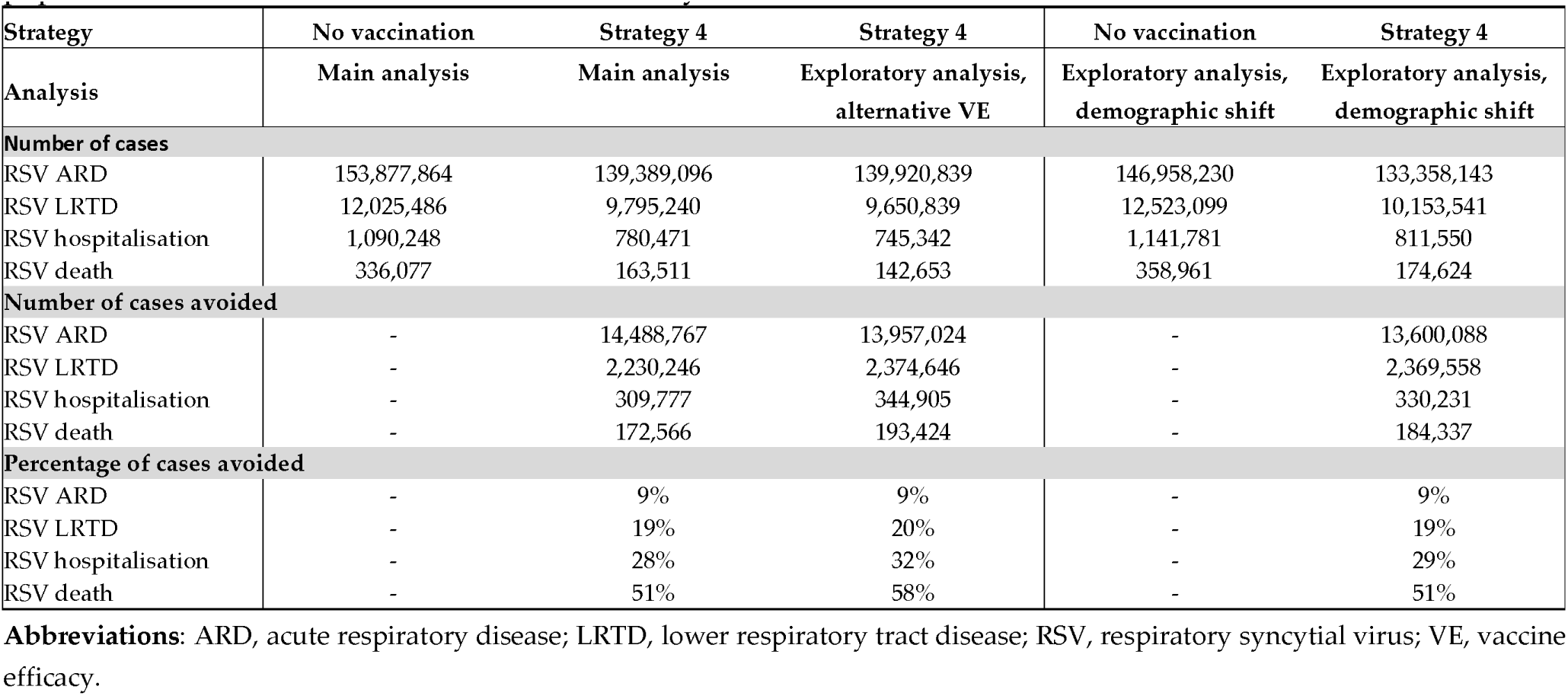
Exploratory analysis: Number of cases, number of cases avoided, and percentage of cases avoided in targeted population, mRNA-1345 vs no vaccination, over 20 years

| Strategy | No vaccination | Strategy 4 | Strategy 4 | No vaccination | Strategy 4 |
| --- | --- | --- | --- | --- | --- |
| Analysis | Main analysis | Main analysis | Exploratory analysis, alternative VE | Exploratory analysis, demographic shift | Exploratory analysis, demographic shift |
| **Number of cases** |  |  |  |  |  |
| RSV ARD | 153,877,864 | 139,389,096 | 139,920,839 | 146,958,230 | 133,358,143 |
| RSV LRTD | 12,025,486 | 9,795,240 | 9,650,839 | 12,523,099 | 10,153,541 |
| RSV hospitalisation | 1,090,248 | 780,471 | 745,342 | 1,141,781 | 811,550 |
| RSV death | 336,077 | 163,511 | 142,653 | 358,961 | 174,624 |
| **Number of cases avoided** |  |  |  |  |  |
| RSV ARD | - | 14,488,767 | 13,957,024 | - | 13,600,088 |
| RSV LRTD | - | 2,230,246 | 2,374,646 | - | 2,369,558 |
| RSV hospitalisation | - | 309,777 | 344,905 | - | 330,231 |
| RSV death | - | 172,566 | 193,424 | - | 184,337 |
| **Percentage of cases avoided** |  |  |  |  |  |
| RSV ARD | - | 9% | 9% | - | 9% |
| RSV LRTD | - | 19% | 20% | - | 19% |
| RSV hospitalisation | - | 28% | 32% | - | 29% |
| RSV death | - | 51% | 58% | - | 51% |

**Abbreviations:** ARD, acute respiratory disease; LRTD, lower respiratory tract disease; RSV, respiratory syncytial virus; VE, vaccine efficacy.

In the scenario applying an alternative assumption on vaccine efficacy, slightly lower number of RSV-ARD cases avoided was expected, while the benefits in terms of RSV-related LRTD, hospitalisations, and deaths were more pronounced than those observed with the main analysis results. These changes indicate that the higher short-term efficacy and/or slower waning, if confirmed, may lead to even greater public health impact than estimated in this analysis, particularly for severe RSV outcomes. In the scenario incorporating demographic shift, a separate simulation of the strategy without vaccination was required, in order to project RSV epidemiology and impact of vaccination under the alternative population structure. In this scenario, population size was assumed to increase faster (as compared to the main analysis), with progressively decreasing proportion of children and increasing proportion of older adults (see also **Supplement 2**). Therefore, in absence of vaccination, the number of RSV infections was expected to decrease slightly, likely reflecting the decreasing proportion of children in the population, while this age group substantially contribute to RSV transmission. However, the number of severe RSV outcomes was projected to be higher compared with the main analysis results, due to a larger proportion of older individuals at high risk for hospitalisation and death. Under such conditions, vaccination against RSV for older adults is expected to provide more pronounced long-term benefits.

## 4. Discussion

Prevention of RSV-related burden in older adults is considered a public health priority in the UK due to increasing recognition of the significant burden in the ageing population and its contribution to seasonal pressure on NHS [17], which also aligns with a government’s health mission emphasizing the importance of proactive approaches to health protection [61,62]. Initial efforts to address this unmet need began in 2024-2025 with a pilot vaccination programme targeting individuals aged 75–80 years [1], with JCVI recommendation recently extended to adults aged 80 years and older, as well as all residents in a care home for older adults [15]. This extension program was based on new evidence, including real-world UK data demonstrating vaccine effectiveness in those aged ≥80 years, increased RSV-related hospitalisations and mortality with advancing age, frequent and severe outbreaks in care homes, and cost-effectiveness analyses showing incrementally greater benefit in the ≥80-year age group compared with those aged 75–79 years. Additional supporting data included the safety profile of RSV vaccines and evidence supporting co-administration with COVID-19 vaccines [15]. Further policy discussions are ongoing to extend the programme, including broader age eligibility, revaccination, and introduction of additional vaccine products [23,24].

This analysis focused on the mRNA-1345 vaccine, developed by Moderna, which has shown robust efficacy and favourable safety in older adults, as demonstrated in the ConquerRSV trial [25-27]. Notably, in the ConquerRSV trial, rare neurological events such as Guillain-Barré syndrome or acute disseminated encephalomyelitis have not been observed following mRNA-1345 administration within the 42-day risk window post-vaccination. While all three RSV products have demonstrated a promising profile, the lack of comparative evidence [1] did not allow for a robust model-based comparison between vaccines. Therefore, mRNA-1345 vaccination strategies were compared to a hypothetical scenario without vaccination.

The results of this analysis confirmed that vaccination with mRNA-1345 could substantially reduce the clinical burden of RSV in older adults, with the magnitude of benefit varying depending on the type of immunisation campaign considered. While the current UK policy targeting individuals aged 75-80 years provides initial protection, its long-term impact remains limited due to the narrow age range and absence of revaccination strategies. It was shown that providing a sustainable protection against RSV over the long-term would require programme designs that incorporate repeated vaccination and broader age eligibility. Vaccination strategies targeting adults aged ≥60 years with regular revaccination intervals demonstrated the greatest reduction in RSV burden, preventing up to 294,000 hospitalisations in the target population over a 20-year timeframe.

In addition to the direct effect among vaccinees, the model projections suggest that RSV vaccination in older adults is also expected to confer indirect protection for individuals outside the target population, resulting in tangible clinical benefits at the total population level, with up to 310,000 hospitalisations prevented over a 20-year timeframe. The largest indirect effects were observed in age cohorts adjacent to the target population, which indicates the potential to prevent RSV-related diseases in individuals who are at increasing risk of severe outcomes, but are not yet eligible for immunisation.

While the settings applied in the main analysis aimed to represent a conservative approach, two exploratory analyses were conducted to illustrate how variations in key assumptions may influence the expected public health impact of mRNA-1345 vaccination. First, if future data confirm a more favourable vaccine effectiveness profile, particularly in terms of higher short-term protection against severe disease outcomes, which could not be fully assessed in the clinical trial due to the limited number of RSV-associated hospitalisations, the actual public health benefit of vaccination against RSV may exceed the estimates presented in this analysis.

The second exploratory scenario investigated the impact of the projected demographic shift, including accelerated ageing and population growth, which are rarely accounted for in modelling studies for RSV, but appear relevant for long-term public health programmes. As might be expected, vaccination against RSV in older adults would provide more pronounced benefits over the long-term, if the real-world population growth and ageing trend is aligned with the current population projections.

The findings of this study should be interpreted considering several strengths and limitations of the modelling approach.

Among the key strengths of this analysis is the application of a well-established dynamic transmission modelling framework, with a final model calibrated and validated using UK-specific demographic and epidemiological data. The model reproduced observed age-specific and seasonal patterns of RSV incidence and hospitalisations well, which supports the credibility of projected public health impact estimates. Capturing the effects of demographic shift strengthens the relevance of this analysis for long-term planning. While rarely incorporated in RSV transmission models, accounting for population dynamics is increasingly important for designing the long-term immunisation strategies. This study contributes to that evidence base, setting the precedent and highlighting the feasibility and policy value of integrating forward-looking demographic assumptions into vaccination impact assessments.

This study also has several limitations, primarily driven by uncertainty in the underlying epidemiological and vaccine-related inputs. While the model was calibrated to reproduce age-specific RSV hospitalisation and RSV-ARD incidence, the real burden of RSV in the UK population remains uncertain, particularly for mortality. For example, a recent UKHSA webinar for health professionals indicated that the reported annual number of RSV-related deaths in individuals ≥65 years in England varies very broadly across different studies, with a more than a 15-fold difference between the lowest and highest of the presented estimates [63]. In this analysis, probability of death given RSV hospitalization was based on the relevant studies by Fleming et al. [46] and Korsten et al. [47], however given the known limitations in RSV death reporting, the projections reported in this study should be interpreted with caution. It should be also noted that granular data on the age distribution of RSV outcomes are limited, which hinders model parametrisation, and likely contributes to uncertainty in age-specific projections of disease burden and clinical impact. Additionally, real-world effectiveness data for mRNA-1345 are not yet available. This highlights the need for dedicated evidence generation activities, including enhanced RSV surveillance and prospective studies, to improve the accuracy of disease burden estimates and support future policy development.

Choice of the recommended vaccination strategies will depend on the national policy objectives, informed by robust evidence on disease burden, vaccine performance, and health system priorities. While this study focused on the epidemiological impact of alternative approaches, a formal cost-effectiveness analysis was beyond its scope, although it will certainly be required to support decision-making. Such analysis should be conducted alongside efforts to strengthen the evidence base on the RSV burden estimates, comparative vaccine performance and key outcomes of interest. This modelling study suggests that strategies combining broader age eligibility (e.g. ≥60 years) with revaccination at regular intervals would be required to achieve a sustainable reduction in RSV burden. Such an approach not only maximises direct protection but also confers indirect benefits across the population. Targeting adults aged ≥60 years is supported by the age-related rise in RSV risk and by broader public health considerations. Individuals in this age range are increasingly vulnerable but may still be economically active and socially engaged, making them a high-impact group for preventive interventions. Expanding the eligible population beyond the currently recommended 75+ group aligns with the UK’s strategic health policy goals, including prevention, reducing hospital burden, and delivering care closer to home [61,64]. A wider vaccination strategy may also improve equity by reaching at-risk individuals earlier in the ageing process, particularly those with comorbidities who are not yet eligible under current guidance.

## 5. Conclusions

Extended RSV vaccination strategies including broader age eligibility and routine revaccination could offer substantial public health benefits in the UK. Targeting adults aged ≥60 years is expected to be particularly efficient in achieving a sustainable reduction in RSV burden and alleviating seasonal pressures on the healthcare system. These findings could provide valuable support for national policy discussions on optimising RSV vaccination strategies in older adults, particularly with regard to target age groups, revaccination schedules, and long-term programme planning.

## Author Contributions

MD, AT, and ZJ: Conceptualization, methodology, data curation, formal analysis, validation, writing—original draft preparation, writing—review and editing, project administration; PO, OB, and KJ: Conceptualization, methodology, validation, writing—review and editing, project administration. All authors have read and agreed to the published version of the manuscript.

## Institutional Review Board Statement

Not applicable.

## Informed Consent Statement

Not applicable.

## Data Availability Statement

The original contributions presented in this study are included in the article/Supplementary Materials, and further inquiries can be directed to the corresponding author.

## Acknowledgments

Medical writing and editorial assistance were provided by Louansha Nandlal, PhD, and Agnieszka Looney, PhD, of MEDiSTRAVA in accordance with Good Publication Practice (GPP 2022) guidelines, funded by Moderna, Inc., and under the direction of the authors.

## 6. Abbreviations

The following abbreviations are used in this manuscript:

ARD: Acute respiratory disease
DTM: Dynamic transmission model
JCVI: Joint Committee on Vaccination and Immunisation
LRTD: Lower respiratory tract disease
mRNA: Messenger ribonucleic acid
NHS: National Health Service
PreF: Prefusion
RSV: Respiratory syncytial virus
UKHSA: United Kingdom Health Security Agency
VE: Vaccine efficacy

