## Supplement for "The Potential Public Health Impact of the mRNA-based Respiratory Syncytial Virus Vaccine, mRNA-1345, Under Extended Vaccination Campaigns Among Older Adults in the United Kingdom: A Modelling Study"

#### S1 Supplement 1: Detailed methodology and inputs

##### *S1.1 Modelling methodology*

###### S1.1.1 Model compartments

While the full model includes compartments for both vaccinated and unvaccinated individuals to capture differential infection risks and disease progression, an additional version excluding vaccination-related compartments was used during model calibration and simulations for strategy without vaccination. This approach minimized computational burden while preserving core transmission dynamics. **Table S1** provides a brief description of the model compartments.

**Table S1.** Description of the epidemiological compartments of the model.

| Compartment | Descriptions |
|---|---|
| **Main compartments** | |
| $\boldsymbol{M}$ | Individuals completely protected from RSV infection due to maternal immunity |
| $\boldsymbol{S_i}$ for $\boldsymbol{i\,\epsilon(1,2,3,4)}$ | Individuals susceptible to RSV infection, before $i$-th infection |
| $\boldsymbol{E_i}$ for $\boldsymbol{i\,\epsilon(1,2,3,4)}$ | Individuals infected by RSV, but not yet infectious (i.e., exposed), who have experienced $i$ infections (including a current one) |
| $\boldsymbol{A_i}$ for $\boldsymbol{i\,\epsilon(1,2,3,4)}$ | Individuals infected by RSV, infectious, and asymptomatic, who have experienced $i$ infections (including a current one) |
| $\boldsymbol{I_i}$ for $\boldsymbol{i\,\epsilon(1,2,3,4)}$ | Individuals infected by RSV, infectious, and symptomatic (with ARD), who have experienced $i$ infections (including a current one) |
| $\boldsymbol{R_i}$ for $\boldsymbol{i\,\epsilon(1,2,3,4)}$ | Individuals recovered from RSV infection, completely protected from a new infection due to natural immunity, who have experienced $i$ previous infections |
| $\boldsymbol{V_4^j}$ for j $\boldsymbol{\epsilon(1,\dots,157)}$ | Vaccinated individuals, who have experienced 3+ previous infections, $j$-th week since vaccination |
| $\boldsymbol{E_{V4}}$ | Vaccinated individuals, infected by RSV, but not yet infectious (i.e., exposed), who have experienced 4+ infections (including a current one) |
| $\boldsymbol{A_{V4}}$ | Vaccinated individuals, infected by RSV, infectious, and asymptomatic, who have experienced 4+ infections (including a current one) |
| $\boldsymbol{I_{V4}}$ | Vaccinated individuals, infected by RSV, infectious, and symptomatic (with ARD), who have experienced 4+ infections (including a current one) |
| **Observational compartments** | |
| $\boldsymbol{INF}$ | Total number of RSV infections (symptomatic and asymptomatic) |
| $\boldsymbol{ARD}$ | Number of ARD cases |
| $\boldsymbol{L}$ | Number of LRTD cases |
| $\boldsymbol{H}$ | Number of hospitalized cases due to LRTD |
| $\boldsymbol{D}$ | Number of fatal cases due to hospitalized LRTD |

**Abbreviations:** ARD, acute respiratory disease; LRTD, lower respiratory tract disease; RSV, respiratory syncytial virus.

**Note:** Each of the model compartments was stratified into 5-year age groups before 85 years, and a group ≥85 years.

###### S1.1.2 Model parameters

Ordinary differential equations (ODE) and additional calculations were parameterized to provide a mathematical notation. A list of parameters used in the model calculations is presented in **Table S2**.

**Table S2.** Parameters used in the differential equations.

| Parameter | Type | Description |
|---|---|---|
| | | **Meta-parameters** |
| $\boldsymbol{M}$ | | Individuals completely protected from RSV infection due to maternal immunity |
| $\boldsymbol{q}$ | Variable | Indicates whether transmission type depends on population density or not (density-dependent transmission or frequency-dependent transmission, 0 or 1 respectively): |
| $\boldsymbol{a}, \boldsymbol{k}$ | Variable | Age group, according to age stratification in the model |
| $\boldsymbol{t}$ | Variable | Time point, in weeks |
| | | **Demographic parameters** |
| $\boldsymbol{N(t)}$ | Calculated within ODE | Total population size at time $t$ |
| $\boldsymbol{N(a,t)}$ | Calculated within ODE | Population size in age group $a$, at time $t$ |
| $\boldsymbol{\xi(a)}$ | Calculated within ODE | Ageing rate from age group $a$ to age group $a+1$ |
| $\boldsymbol{c_{ak}}$ | Input | Number of contacts between a person from age group $a$ and a person from age group $k$ |
| $\boldsymbol{\mu_{pop}(a)}$ | Input | Rate of change for the population size, including general mortality and migration in population, for age group $a$ |
| $\boldsymbol{birth\_rate}$ | Input | The rate at which individuals enter the model |
| | | **Epidemiological parameters required to estimate the force of infection (FOI)** |
| $\boldsymbol{\lambda(a,t)}$ | Calculated within ODE | FOI in age group $a$, at time $t$ |
| $\boldsymbol{\beta_{a,k}}$ | Calculated within ODE | Transmission rate from age group $k$ to age group $a$ |
| $\boldsymbol{b}$ | Estimated in calibration | Transmission probability (transmission probability from infectious to susceptible individual) |
| $\boldsymbol{s_{amp}}$ | Estimated in calibration | Seasonality parameter, amplitude of sinusoidal function |
| $\boldsymbol{s_{shift}}$ | Estimated in calibration | Seasonality parameter, horizontal shift of sinusoidal function |
| $\boldsymbol{\psi_i}$ for $\boldsymbol{i \in (1,2,3,4)}$ | Input | Relative infectiousness of $i$-th infection, equal to 1 for $i$=1 |
| | | **Other epidemiological parameters** |
| $\boldsymbol{\varepsilon}$ | Input | Rate of becoming infectious |
| $\boldsymbol{\gamma_i}$ for $\boldsymbol{i \in (1,2,3,4)}$ | Input | Recovery rate from $i$-th infection |
| $\boldsymbol{\sigma_i}$ for $\boldsymbol{i \in (1,2,3)}$ | Input | Relative risk of re-infection, after $i$-th infection |
| $\boldsymbol{\rho(a)}$ | Input | Proportion of infections that are asymptomatic, for age group $a$ |
| $\boldsymbol{\kappa_i(a)}$ for $\boldsymbol{i \in (1,2,3,4)}$ | Input | Proportion of ARD infections that are LRTDs, for age group $a$, for $i$-th infection |
| $\boldsymbol{\delta_i(a)}$ for $\boldsymbol{i \in (1,2,3,4)}$ | Input | Proportion of hospitalisations due to LRTDs, for age group $a$, for $i$-th infection |
| $\boldsymbol{\mu(a)}$ | Input | Proportion of deaths due to hospitalized RSV, for age group $a$ |
| $\boldsymbol{\eta_m}$ | Input | Waning rate of maternal immunity |
| $\boldsymbol{\eta_i}$ for $\boldsymbol{i \in (1,2,3,4)}$ | Input | Waning rate of post-infection immunity, for $i$-th infection |
| | | **Vaccine related parameters** |
| $\boldsymbol{\chi_{j,t_{RV}}}$ | Variable | Parameter which indicates whether the individual in $V_4^j$ compartment is revaccinated (1) or not (0),<br>equal 1 when j > $t_{RV}$, 0 when j ≤ $t_{RV}$, in $j$-th week since last vaccination |
| $\boldsymbol{t_{RV}}$ | Input | Number of weeks after which individual with previous vaccination can be revaccinated |
| $\boldsymbol{\theta(a,t)}$ | Input | Vaccination coverage for age group $a$ at time $t$ |
| $\boldsymbol{\sigma_4^j}$ | Input | Relative risk of RSV infection in vaccinated versus unvaccinated individuals, in $j$-th week since vaccination |
| $\boldsymbol{\sigma_5^j}$ | Input | Relative risk of LRTD given ARD in vaccinated versus unvaccinated individuals, in $j$-th week since vaccination |
| $\boldsymbol{\sigma_6^j}$ | Input | Relative risk of hospitalisation given LRTD in vaccinated versus unvaccinated individuals, in $j$-th week since vaccination |
| $\boldsymbol{\gamma_{V4}}$ | Input | Recovery rate from 4+ infection for a vaccinated individual |

**Abbreviations:** ARD, acute respiratory disease; FOI, force of infection; LRTD, lower respiratory tract disease; ODE, ordinary differential equations; RSV, respiratory syncytial virus.

##### S1.1.3 Differential equations

The ODEs used in the model are provided below, for age group $a$ are at time $t$:

**Compartments for individuals with no previous respiratory syncytial virus (RSV) infection**

$$\frac{dM(a,t)}{dt} = birth\_rate \cdot N(t) - M(a,t) \cdot \left(\eta_m + \mu_{pop}(a)\right) - M(a,t) \cdot \xi(a) + M(a-1,t) \cdot \xi(a-1)$$

$$\frac{dS_1(a,t)}{dt} = M(a,t) \cdot \eta_m - S_1(a,t) \cdot \left(\lambda(a,t) + \mu_{pop}(a)\right) - S_1(a,t) \cdot \xi(a) + S_1(a-1,t) \cdot \xi(a-1)$$

$$\frac{dE_1(a,t)}{dt} = S_1(a,t) \cdot \lambda(a,t) - E_1(a,t) \cdot \left(\varepsilon + \mu_{pop}(a)\right) - E_1(a,t) \cdot \xi(a) + E_1(a-1,t) \cdot \xi(a-1)$$

$$\frac{dA_1(a,t)}{dt} = E_1(a,t) \cdot \varepsilon \cdot \rho(a) - A_1(a,t) \cdot \left(\gamma_1 + \mu_{pop}(a)\right) - A_1(a,t) \cdot \xi(a) + A_1(a-1,t) \cdot \xi(a-1)$$

$$\frac{dI_1(a,t)}{dt} = E_1(a,t) \cdot \varepsilon \cdot \left(1 - \rho(a)\right) - I_1(a,t) \cdot \left(\gamma_1 + \mu_{pop}(a)\right) - I_1(a,t) \cdot \xi(a) + I_1(a-1,t) \cdot \xi(a-1)$$

$$\frac{dR_1(a,t)}{dt} = \left(A_1(a,t) + I_1(a,t)\right) \cdot \gamma_1 - R_1(a,t) \cdot \left(\eta_1 + \mu_{pop}(a)\right) - R_1(a,t) \cdot \xi(a) + R_1(a-1,t) \cdot \xi(a-1)$$

**Compartments for individuals with one previous RSV infection**

$$\frac{dS_2(a,t)}{dt} = R_1(a,t) \cdot \eta_1 - S_2(a,t) \cdot \left(\lambda(a,t) \cdot \sigma_1 + \mu_{pop}(a)\right) - S_2(a,t) \cdot \xi(a) + S_2(a-1,t) \cdot \xi(a-1)$$

$$\frac{dE_2(a,t)}{dt} = S_2(a,t) \cdot \lambda(a,t) \cdot \sigma_1 - E_2(a,t) \cdot \left(\varepsilon + \mu_{pop}(a)\right) - E_2(a,t) \cdot \xi(a) + E_2(a-1,t) \cdot \xi(a-1)$$

$$\frac{dA_2(a,t)}{dt} = E_2(a,t) \cdot \varepsilon \cdot \rho(a) - A_2(a,t) \cdot \left(\gamma_2 + \mu_{pop}(a)\right) - A_2(a,t) \cdot \xi(a) + A_2(a-1,t) \cdot \xi(a-1)$$

$$\frac{dI_2(a,t)}{dt} = E_2(a,t) \cdot \varepsilon \cdot \left(1 - \rho(a)\right) - I_2(a,t) \cdot \left(\gamma_2 + \mu_{pop}(a)\right) - I_2(a,t) \cdot \xi(a) + I_2(a-1,t) \cdot \xi(a-1)$$

$$\frac{dR_2(a,t)}{dt} = \left(A_2(a,t) + I_2(a,t)\right) \cdot \gamma_2 - R_2(a,t) \cdot \left(\eta_2 + \mu_{pop}(a)\right) - R_2(a,t) \cdot \xi(a) + R_2(a-1,t) \cdot \xi(a-1)$$

**Compartments for individuals with two previous RSV infections**

$$\frac{dS_3(a,t)}{dt} = R_2(a,t) \cdot \eta_2 - S_3(a,t) \cdot \left(\lambda(a,t) \cdot \sigma_2 + \mu_{pop}(a)\right) - S_3(a,t) \cdot \xi(a) + S_3(a-1,t) \cdot \xi(a-1)$$

$$\frac{dE_3(a,t)}{dt} = S_3(a,t) \cdot \lambda(a,t) \cdot \sigma_2 - E_3(a,t) \cdot \left(\varepsilon + \mu_{pop}(a)\right) - E_3(a,t) \cdot \xi(a) + E_3(a-1,t) \cdot \xi(a-1)$$

$$\frac{dA_3(a,t)}{dt} = E_3(a,t) \cdot \varepsilon \cdot \rho(a) - A_3(a,t) \cdot \left(\gamma_3 + \mu_{pop}(a)\right) - A_3(a,t) \cdot \xi(a) + A_3(a-1,t) \cdot \xi(a-1)$$

$$\frac{dI_3(a,t)}{dt} = E_3(a,t) \cdot \varepsilon \cdot \left(1 - \rho(a)\right) - I_3(a,t) \cdot \left(\gamma_3 + \mu_{pop}(a)\right) - I_3(a,t) \cdot \xi(a) + I_3(a-1,t) \cdot \xi(a-1)$$

$$\frac{dR_3(a,t)}{dt} = \left(A_3(a,t) + I_3(a,t)\right) \cdot \gamma_3 - R_3(a,t) \cdot \left(\eta_3 + \mu_{pop}(a)\right) - R_3(a,t) \cdot \xi(a) + R_3(a-1,t) \cdot \xi(a-1)$$

**Compartments for individuals with three or more previous RSV infections**

$$\frac{dS_4(a,t)}{dt} = R_3(a,t) \cdot \eta_3 + R_4(a,t) \cdot \eta_4 - S_4(a,t) \cdot \left(\lambda(a,t) \cdot \sigma_3 - \theta(a,t) + \mu_{pop}(a)\right) - S_4(a,t) \cdot \xi(a) + S_4(a-1,t) \cdot \xi(a-1)$$

$$\frac{dV_4^1(a,t)}{dt} = S_4(a,t) \cdot \theta(a,t) + \sum_{j=t_{RV}+1}^{157} V_4^j(a,t) \cdot \theta(a,t) - V_4^1(a,t)$$

$$\begin{aligned}\frac{dV_4^j(a,t)}{dt} = {} & V_4^{j-1}(a,t) \cdot \left(1 - \lambda(a,t) \cdot \sigma_3 \cdot \sigma_4^{j-1} - \chi_{j-1,t_{RV}} \cdot \theta(a,t) - \mu_{pop}(a)\right) - V_4^{j-1}(a,t) \\ & \cdot \left(1 - \lambda(a,t) \cdot \sigma_3 \cdot \sigma_4^{j-1} - \chi_{j-1,t_{RV}} \cdot \theta(a,t) - \mu_{pop}(a)\right) \cdot \xi(a) + V_4^{j-1}(a-1,t) \\ & \cdot \left(1 - \lambda(a-1,t) \cdot \sigma_3 \cdot \sigma_4^{j-1} - \chi_{j-1,t_{RV}} \cdot \theta(a-1,t) - \mu_{pop}(a-1)\right) \cdot \xi(a-1) - V_4^j(a,t), 1 < j \\ & < 157\end{aligned}$$

$$\frac{dV_4^{157}(a,t)}{dt} = V_4^{156}(a,t)\cdot\Big(1-\lambda(a,t)\cdot\sigma_3\cdot\sigma_4^{156}-\chi_{156,t_{RV}}\cdot\theta(a,t)-\mu_{pop}(a)\Big)-V_4^{156}(a,t)$$
$$\cdot\Big(1-\lambda(a,t)\cdot\sigma_3\cdot\sigma_4^{156}-\chi_{156,t_{RV}}\cdot\theta(a,t)-\mu_{pop}(a)\Big)\cdot\xi(a)+V_4^{156}(a-1,t)$$
$$\cdot\Big(1-\lambda(a-1,t)\cdot\sigma_3\cdot\sigma_4^{156}-\chi_{156,t_{RV}}\cdot\theta(a-1,t)-\mu_{pop}(a-1)\Big)\cdot\xi(a-1)-V_4^{157}(a,t)$$
$$\cdot\Big(\lambda(a,t)\cdot\sigma_3\cdot\sigma_4^{157}+\chi_{157,t_{RV}}\cdot\theta(a,t)+\mu_{pop}(a)\Big)-V_4^{157}(a,t)\cdot\xi(a)+V_4^{157}(a-1,t)\cdot\,\xi(a-1)$$

$$\frac{dE_4(a,t)}{dt} = S_4(a,t)\cdot\lambda(a,t)\cdot\sigma_3-E_4(a,t)\cdot\big(\varepsilon+\mu_{pop}(a)\big)-E_4(a,t)\cdot\xi(a)+E_4(a-1,t)\cdot\,\xi(a-1)$$

$$\frac{dE_{V4}(a,t)}{dt} = \sum_{j=1}^{157}V_4^j(a,t)\cdot\lambda(a,t)\cdot\sigma_3\cdot\sigma_4^j-E_{V4}(a,t)\cdot\Big(\varepsilon+\mu_{pop}(a)\Big)-E_{V4}(a,t)\cdot\xi(a)+E_{V4}(a-1,t)\cdot\,\xi(a-1)$$

$$\frac{dA_4(a,t)}{dt} = E_4(a,t)\cdot\varepsilon\cdot\rho(a)-A_4(a,t)\cdot\big(\gamma_4+\mu_{pop}(a)\big)-A_4(a,t)\cdot\xi(a)+A_4(a-1,t)\cdot\,\xi(a-1)$$

$$\frac{dA_{V4}(a,t)}{dt} = E_{V4}(a,t)\cdot\varepsilon\cdot\rho(a)-A_{V4}(a,t)\cdot\Big(\gamma_{V4}+\mu_{pop}(a)\Big)-A_{V4}(a,t)\cdot\xi(a)+A_{V4}(a-1,t)\cdot\,\xi(a-1)$$

$$\frac{dI_4(a,t)}{dt} = E_4(a,t)\cdot\varepsilon\cdot\big(1-\rho(a)\big)-I_4(a,t)\cdot\Big(\gamma_4+\mu_{pop}(a)\Big)-I_4(a,t)\cdot\xi(a)+I_4(a-1,t)\cdot\,\xi(a-1)$$

$$\frac{dI_{V4}(a,t)}{dt} = E_{V4}(a,t)\cdot\varepsilon\cdot\big(1-\rho(a)\big)-I_{V4}(a,t)\cdot\Big(\gamma_{V4}+\mu_{pop}(a)\Big)-I_{V4}(a,t)\cdot\xi(a)+I_{V4}(a-1,t)\cdot\,\xi(a-1)$$

$$\frac{dR_4(a,t)}{dt} = \big(A_4(a,t)+I_4(a,t)\big)\cdot\gamma_4+\big(A_{V4}(a,t)+I_{V4}(a,t)\big)\cdot\gamma_{V4}-R_4(a,t)\cdot\Big(\eta_4+\mu_{pop}(a)\Big)-R_4(a,t)\cdot\xi(a)$$
$$+R_4(a-1,t)\cdot\,\xi(a-1)$$

##### S1.1.4 Force of infection

The force of infection defines the risk of acquiring RSV for a susceptible individual, i.e., a transition from compartment $S$ (susceptible) to compartment $E$ (exposed).

It was assumed that the probability of transmission is seasonal, and it was modelled using a cosine function.

The chance of being infected for susceptible individuals in the age group $a$ depends on their contacts with infectious individuals in population. The transmission rate from age group $k$ to age group $a$, denoted $\beta_{a,k}$, is estimated using the age-specific number of contacts between age group $a$ and k per time step ($c_{a,k}$) and the probability of transmission given contact between an infectious and a susceptible individual ($b$). Parameter $q$ indicates whether transmission depends or not on the population size (q equal to 0 or 1, respectively), with q assumed equal to 1 for the current model settings:

$$\beta_{a,k}(t) = \frac{b\cdot c_{a,k}}{N(t)^{1-q}},$$

where $N(t)$ denotes the total population size at time $t$.

Thus, the force of infection ($\lambda$) for susceptible individual in age group $a$, at time $t$, was calculated as follows:

$$\lambda(a,t) = \left(1+s_{amp}\cdot\exp\left(cos\left(\frac{2\cdot\pi\cdot\big(t-s_{shift}\cdot 52\big)}{52}\right)\right)\right)\cdot\sum_k\left(\beta_{a,k}\cdot\frac{\sum_{i=1}^{4}\psi_i\cdot\big(A_i(k,t)+I_i(k,t)\big)}{N(k,t)}\right)$$

where $s_{amp}$ is the seasonality amplitude, $s_{shift}$ is the seasonal shift, $A_i(k,t)$ and $I_i(k,t)$ is the number of asymptomatic and symptomatic infectious individuals in age group $k$ during their $i$-th infection at time $t$, $\psi_i$ is the relative infectiousness of the $i$-th infection; and $N(k,t)$ is population size in age group $k$ at time $t$.

##### S1.1.5 Ageing

The model accounted for ageing at weekly intervals. While all the individuals are undergoing an ageing process, only some of them transition to another age-defined compartment due to ageing. Thus, individuals who transition to another compartment due to ageing represent only a proportion of an age group, at the upper bound of age, who complete a full year of life over a given week. The ageing rate for the age group $a$ was estimated as:

$$\xi(a) = \frac{1}{(a^{hight} - a^{low} + 1) \cdot 52},$$

where $a^{low}$ and $a^{hight}$ are the lower and upper bounds of an age group $a$, respectively, and $52$ is the number of weeks in a year.

S1.1.6 Infection consequences

The modelling approach considered that RSV infection can be symptomatic (acute respiratory disease [ARD]), which can progress to lower respiratory disease (LRTD), that LRTD can be associated with hospitalisation, and that hospitalized LRTD cases can be fatal. The number of infection-related events (RSV infections, ARD, LRTD, hospitalisations and RSV-related deaths) was calculated outside the ODE, within the observational compartments INF, ARD, L, H and D, respectively, using the number of transitions from compartments S to E in the previous time step, and respective severity parameters.

**The number of incident RSV infections (asymptomatic and symptomatic)** was estimated using the number of transitions into the compartment $E$ in the previous time point:

$$INF(a,t) = \sum_{i=1}^{4} INF_i(a,t) + INF_V(a,t) = \sum_{i=1}^{4} INF_i(a,t) + \sum_{j=1}^{157} INF_V^j(a,t)$$

$$= S_1(a,t-1) \cdot \lambda(a,t-1) + \sum_{i=2}^{4} S_i(a,t-1) \cdot \lambda(a,t-1) \cdot \sigma_{i-1}$$

$$+ \sum_{j=1}^{157} V_4^j(a,t-1) \cdot \lambda(a,t-1) \cdot \sigma_3 \cdot \sigma_4^j,$$

where $INF_i(a,t)$ is the number of RSV infections, which are $i$-th infection, for age group $a$, at time $t$, in unvaccinated individuals, $INF_V^j(a,t)$ is the number of RSV infections, for age group $a$, at time $t$, in vaccinated individuals in $j$-th week since vaccination, $S_i(a,t-1)$ is the number of individuals susceptible to RSV infection, for whom the potential infection will be the $i$-th previous infection, $\lambda(a,t-1)$ is a force of infection for age group a, at time $t-1$, $\sigma_i$ is a relative risk of re-infection, after $i$-th infection, $\sigma_4^j$ is a relative risk of RSV infection in vaccinated versus unvaccinated individuals, in $j$-th week since vaccination.

The number of ARD infections (ARD) was estimated using the number of incident RSV infections multiplied by the proportion of symptomatic infections among all infections.

$$ARD(a,t) = \sum_{i=1}^{4} ARD_i(a,t) + ARD_V(a,t) = \sum_{i=1}^{4} ARD_i(a,t) + \sum_{j=1}^{157} ARD_V^j(a,t)$$

$$= \sum_{i=1}^{4} INF_i(a,t) \cdot (1-\rho(a)) + \sum_{j=1}^{157} INF_V^j(a,t) \cdot (1-\rho(a)),$$

where $ARD_i(a,t)$ is the number of ARD infections, which are $i$-th infection, for age group $a$, at time $t$, in unvaccinated individuals, $ARD_V^j(a,t)$ is the number of ARDSV infections, for age group $a$, at time $t$, in vaccinated individuals in $j$-th week since vaccination, $\sigma_i$ is a relative risk of re-infection, after $i$-th infection, $\rho(a)$ is a proportion of infections that are asymptomatic, for age group $a$.

**The number of LRTD infections (L)** was estimated using the number of incident ARD, multiplied by the proportion of ARD infections that are LRTD. For protected individuals, this number was adjusted by the relative risk of LRTD given ARD in vaccinated versus unvaccinated individuals ($\sigma_5^j$).

$$L(a,t) = \sum_{i=1}^{4} L_i(a,t) + L_V(a,t) = \sum_{i=1}^{4} L_i(a,t) + \sum_{j=1}^{157} L_V^j(a,t) = \sum_{i=1}^{4} ARD_i(a,t) \cdot \kappa_i(a) + \sum_{j=1}^{157} ARD_V^j(a,t) \cdot \kappa_4(a) \cdot \sigma_5^j,$$

where $L_i(a,t)$ is the number of LRTD cases, which are $i$-th infection, for age group $a$, at time $t$, in unvaccinated individuals, $LRTD_V(a,t)$ is the number of LRTD cases, for age group $a$, at time $t$, in vaccinated individuals, $\kappa_i(a)$ is the proportion of ARD infections that are LRTDs, for age group $a$, for $i$-th infection, $\sigma_5^j$ is a relative risk of LRTD given ARD in vaccinated versus unvaccinated individuals, in $j$-th week since vaccination.

**The number of hospitalized cases (H)** was estimated using the number of incident LRTD infections, multiplied by the proportion of hospitalisations due to LRTDs. For protected individuals, this number was adjusted by the relative risk of hospitalisation given LRTD in vaccinated versus unvaccinated individuals ($\sigma_6^j$).

$$H(a,t) = \sum_{i=1}^{4} H_i(a,t) + H_V(a,t) = \sum_{i=1}^{4} H_i(a,t) + \sum_{j=1}^{157} H_V^j(a,t) = \sum_{i=1}^{4} L_i(a,t) \cdot \delta_i(a) + \sum_{j=1}^{157} L_V^j(a,t) \cdot \delta_4(a) \cdot \sigma_6^j,$$

where $H_i(a,t)$ is the number of hospitalised LRTD cases, which are $i$-th infection, for age group $a$, at time $t$, in unvaccinated individuals, $H_V(a,t)$ is the number of hospitalized LRTD cases for age group $a$, at time $t$, in vaccinated individuals, $\delta_i(a)$ is a proportion of hospitalisations due to LRTD, for age group $a$, for $i$-th infection, $\sigma_6^j$ is a relative risk of hospitalisation given LRTD in vaccinated versus unvaccinated individuals, in $j$-th week since vaccination.

**The number of RSV-related deaths (D)** was estimated using the number of incident hospitalized LRTD infections, multiplied by the proportion of deaths due to hospitalized LRTD.

$$D(a,t) = \sum_{i=1}^{4} D_i(a,t) + D_V(a,t) = \sum_{i=1}^{4} H_i(a,t) \cdot \mu(a) + H_V(a,t) \cdot \mu(a),$$

where $D_i(a,t)$ is the number of RSV-related deaths due to $i$-th infection, for age group $a$, at time $t$, $\mu(a)$ is the proportion of deaths due to hospitalized LRTD, for age group $a$.

#### *S1.2 Model inputs*

##### S1.2.1 Demographic inputs

Demographic calibration was informed by data on population size by age group for 1994 [1], 2023 [2], and, for exploratory analysis, 2030 [3]. (**Table S3**).

**Table S3**. Population size reported in 1994 and 2023, and projected for 2030.

| Age group | 1994 | 2023 | 2030 |
|---|---|---|---|
| **0-4 years** | 3,852,159 | 3,572,007 | 3,460,590 |
| **5-9 years** | 3,806,147 | 3,925,921 | 3,569,885 |
| **10-14 years** | 3,633,584 | 4,150,287 | 3,925,960 |
| **15-19 years** | 3,386,905 | 4,011,468 | 4,263,029 |
| **20-24 years** | 4,073,944 | 4,097,542 | 4,623,042 |
| **25-29 years** | 4,600,350 | 4,427,747 | 4,696,704 |
| **30-34 years** | 4,522,350 | 4,700,198 | 4,956,393 |
| **35-39 years** | 3,971,309 | 4,636,593 | 5,061,444 |
| **40-44 years** | 3,782,421 | 4,446,226 | 4,950,903 |
| **45-49 years** | 4,033,741 | 4,043,242 | 4,595,621 |
| **50-54 years** | 3,232,360 | 4,522,878 | 4,173,181 |

| Age group | 1994 | 2023 | 2030 |
|---|---|---|---|
| **55-59 years** | 2,998,630 | 4,625,265 | 4,278,284 |
| **60-64 years** | 2,814,061 | 4,181,669 | 4,468,734 |
| **65-69 years** | 2,670,877 | 3,489,709 | 4,211,345 |
| **70-74 years** | 2,556,347 | 3,120,459 | 3,429,888 |
| **75-79 years** | 1,653,458 | 2,843,315 | 2,746,151 |
| **80-84 years** | 1,299,406 | 1,763,312 | 2,409,715 |
| **85-100 years** | 974,096 | 1,707,371 | 2,059,119 |
| **Total** | **57,862,145** | **68,265,209** | **71,879,988** |

Birth rates presented in **Table S4** were estimated from the data reported by the Office for National Statistics: the reported and projected number of births by year was divided by the total population size for the respective year [2-6].

**Table S4.** Birth rates, 1994-2054.

| Year | Birth rate | Year | Birth rate | Year | Birth rate |
|---|---|---|---|---|---|
| **1994** | 0.01297 | **2015** | 0.01194 | **2036** | 0.00948 |
| **1995** | 0.01261 | **2016** | 0.01180 | **2037** | 0.00955 |
| **1996** | 0.01261 | **2017** | 0.01143 | **2038** | 0.00962 |
| **1997** | 0.01246 | **2018** | 0.01101 | **2039** | 0.00970 |
| **1998** | 0.01226 | **2019** | 0.01067 | **2040** | 0.00977 |
| **1999** | 0.01193 | **2020** | 0.01016 | **2041** | 0.00983 |
| **2000** | 0.01153 | **2021** | 0.00998 | **2042** | 0.00988 |
| **2001** | 0.01132 | **2022** | 0.00979 | **2043** | 0.00991 |
| **2002** | 0.01127 | **2023** | 0.00972 | **2044** | 0.00993 |
| **2003** | 0.01166 | **2024** | 0.00975 | **2045** | 0.00992 |
| **2004** | 0.01194 | **2025** | 0.00973 | **2046** | 0.00989 |
| **2005** | 0.01196 | **2026** | 0.00967 | **2047** | 0.00984 |
| **2006** | 0.01231 | **2027** | 0.00961 | **2048** | 0.00978 |
| **2007** | 0.01259 | **2028** | 0.00956 | **2049** | 0.00970 |
| **2008** | 0.01285 | **2029** | 0.00951 | **2050** | 0.00962 |
| **2009** | 0.01269 | **2030** | 0.00948 | **2051** | 0.00953 |
| **2010** | 0.01286 | **2031** | 0.00945 | **2052** | 0.00943 |
| **2011** | 0.01276 | **2032** | 0.00944 | **2053** | 0.00934 |
| **2012** | 0.01276 | **2033** | 0.00943 | **2054** | 0.00925 |
| **2013** | 0.01215 | **2034** | 0.00943 | - | - |
| **2014** | 0.01202 | **2035** | 0.00944 | - | - |

**Table S5** presents calibrated rate of population change by age group used in the model.

**Table S5.** Calibrated rate of population change, by age group.

| Age group | Rate of population change, main analysis | Rate of population change, exploratory analysis (demographic shift) |
|---|---|---|
| **0-4 years** | 0.000036 | 0.000072 |
| **5-9 years** | -0.000197 | -0.000004 |
| **10-14 years** | -0.000063 | -0.000251 |
| **15-19 years** | -0.000076 | -0.000208 |
| **20-24 years** | -0.000048 | -0.000281 |
| **25-29 years** | -0.000324 | -0.000118 |
| **30-34 years** | -0.000323 | -0.000300 |
| **35-39 years** | 0.000070 | -0.000094 |
| **40-44 years** | 0.000063 | -0.000027 |
| **45-49 years** | 0.000428 | 0.000212 |

| | | |
|---|---|---|
| **50-54 years** | -0.000496 | 0.000405 |
| **55-59 years** | -0.000143 | -0.000210 |
| **60-64 years** | 0.000273 | -0.000275 |
| **65-69 years** | 0.000661 | 0.000067 |
| **70-74 years** | 0.000226 | 0.000828 |
| **75-79 years** | 0.000211 | 0.000769 |
| **80-84 years** | 0.002027 | 0.000316 |
| **85-100 years** | 0.003756 | 0.004337 |

##### S1.2.2 Contact matrix

The total number of physical and conversational contacts between different age groups by week was calculated using data from the POLYMOD study [7] and the R package socialmixr [8]. The UK population structure from the year of the POLYMOD study (2005) was utilized in the socialmixr package to ensure symmetry in the contact matrix. The matrix was constructed with the following assumptions:

- Symmetry of contacts (not symmetry of the matrix object).
- In the absence of age-stratified contact data for individuals aged 75 years and older, the contact pattern estimated for the ≥70 year-olds was applied uniformly across the 70–74, 75–79, 80–84, and ≥85-year age groups (i.e., equal by row, distributed by column).
- Social contacts data were weighted by the day of the week to consider different number of contacts in week days and weekends.
- Social contacts data were weighted based on the age distribution of the population.

The obtained contact matrix is presented in **Figure S1Error! Reference source not found.**.

| Age group, y | 0-4 | 5-9 | 10-14 | 15-19 | 20-24 | 25-29 | 30-34 | 35-39 | 40-44 | 45-49 | 50-54 | 55-59 | 60-64 | 65-69 | 70-74 | 75-79 | 80-84 | 85-100 |
|---|---|---|---|---|---|---|---|---|---|---|---|---|---|---|---|---|---|---|
| **0-4** | 13.1 | 5.8 | 3.3 | 2.1 | 3.7 | 5.8 | 6.2 | 7.9 | 3.1 | 2.1 | 2.5 | 2.2 | 1.5 | 0.7 | 0.5 | 0.4 | 0.3 | 0.3 |
| **5-9** | 5.6 | 46.6 | 8.4 | 4.2 | 3.8 | 5.1 | 7.4 | 10.8 | 7.8 | 2.7 | 2.4 | 1.8 | 2.3 | 1.9 | 0.9 | 0.7 | 0.5 | 0.4 |
| **10-14** | 2.9 | 7.8 | 49.3 | 9.0 | 1.8 | 2.6 | 4.0 | 7.8 | 8.5 | 5.2 | 3.0 | 2.5 | 1.4 | 1.4 | 1.1 | 0.9 | 0.7 | 0.6 |
| **15-19** | 1.8 | 3.8 | 8.7 | 45.5 | 8.1 | 5.0 | 3.3 | 6.2 | 6.5 | 6.3 | 4.1 | 2.2 | 1.4 | 2.1 | 1.6 | 1.3 | 1.0 | 0.8 |
| **20-24** | 3.3 | 3.4 | 1.8 | 8.2 | 18.2 | 9.4 | 6.0 | 5.2 | 5.4 | 7.4 | 4.2 | 3.3 | 2.3 | 1.8 | 1.1 | 0.9 | 0.7 | 0.5 |
| **25-29** | 5.4 | 4.8 | 2.6 | 5.3 | 9.8 | 12.5 | 7.9 | 6.4 | 6.2 | 6.4 | 6.0 | 5.1 | 3.4 | 2.8 | 0.8 | 0.7 | 0.5 | 0.4 |
| **30-34** | 5.1 | 6.4 | 3.7 | 3.1 | 5.6 | 7.1 | 11.3 | 8.9 | 7.0 | 5.3 | 4.5 | 4.6 | 3.2 | 1.8 | 0.8 | 0.7 | 0.5 | 0.4 |
| **35-39** | 5.8 | 8.2 | 6.4 | 5.2 | 4.3 | 5.1 | 7.9 | 10.6 | 8.6 | 6.2 | 4.6 | 3.9 | 3.9 | 3.4 | 1.0 | 0.8 | 0.6 | 0.5 |
| **40-44** | 2.3 | 6.0 | 7.0 | 5.5 | 4.5 | 5.0 | 6.3 | 8.7 | 9.6 | 8.5 | 5.0 | 4.1 | 3.7 | 2.6 | 2.0 | 1.7 | 1.3 | 1.0 |
| **45-49** | 1.8 | 2.4 | 5.0 | 6.3 | 7.2 | 6.1 | 5.5 | 7.3 | 9.9 | 13.2 | 5.6 | 5.0 | 3.8 | 2.0 | 1.7 | 1.4 | 1.1 | 0.9 |
| **50-54** | 2.4 | 2.3 | 3.2 | 4.5 | 4.5 | 6.2 | 5.1 | 6.0 | 6.4 | 6.2 | 5.5 | 7.0 | 3.2 | 2.1 | 2.3 | 1.9 | 1.5 | 1.2 |
| **55-59** | 1.9 | 1.6 | 2.4 | 2.3 | 3.3 | 4.9 | 4.9 | 4.7 | 4.9 | 5.1 | 6.5 | 9.0 | 5.8 | 3.0 | 1.8 | 1.5 | 1.1 | 0.9 |
| **60-64** | 1.7 | 2.6 | 1.7 | 1.7 | 2.9 | 4.1 | 4.3 | 5.8 | 5.5 | 4.9 | 3.7 | 7.3 | 4.6 | 4.6 | 2.1 | 1.8 | 1.3 | 1.1 |
| **65-69** | 1.0 | 2.4 | 1.9 | 3.1 | 2.7 | 3.9 | 2.8 | 5.9 | 4.5 | 3.0 | 2.8 | 4.4 | 5.3 | 5.3 | 2.9 | 2.4 | 1.8 | 1.4 |
| **70-74** | 0.8 | 1.3 | 1.8 | 2.7 | 1.8 | 1.3 | 1.5 | 1.9 | 4.0 | 2.9 | 3.6 | 2.9 | 2.8 | 3.3 | 3.4 | 2.8 | 2.2 | 1.7 |
| **75-79** | 0.8 | 1.3 | 1.8 | 2.7 | 1.8 | 1.3 | 1.5 | 1.9 | 4.0 | 2.9 | 3.6 | 2.9 | 2.8 | 3.3 | 3.4 | 2.8 | 2.2 | 1.7 |
| **80-84** | 0.8 | 1.3 | 1.8 | 2.7 | 1.8 | 1.3 | 1.5 | 1.9 | 4.0 | 2.9 | 3.6 | 2.9 | 2.8 | 3.3 | 3.4 | 2.8 | 2.2 | 1.7 |
| **85-100** | 0.8 | 1.3 | 1.8 | 2.7 | 1.8 | 1.3 | 1.5 | 1.9 | 4.0 | 2.9 | 3.6 | 2.9 | 2.8 | 3.3 | 3.4 | 2.8 | 2.2 | 1.7 |

**Figure S1**. Age-stratified weekly contact matrix (heatmap), with colour intensity indicating the average number of contacts between age groups.

**Abbreviations:** y, years.

##### S1.2.3 Epidemiological inputs

The hospitalisation rate per 100,000 individuals was derived from national surveillance reports [9] and adjusted for underreporting [10] in age groups 0–64 years, while for individuals aged 65 years and older, the rate was based on estimates from Osei-Yeboah et al. [11] (**Table S6**).

**Table S6.** Hospitalisation rate, per 100,000 individuals (age group).

| Age group | Calibration target | Source |
|---|---|---|
| **0-4 years** | 609.544 | |
| **5-14 years** | 20.564 | |
| **15-44 years** | 7.764 | National surveillance reports [30] adjusted for underreporting [10] |
| **45-54 years** | 8.731 | |
| **55-64 years** | 21.780 | |
| **65-74 years** | 77.000 | |
| **75-84 years** | 231.000 | Osei-Yeboah et al. 2023 [11] |
| **85+ years** | 395.000 | |

The weekly hospitalisation rate per 100,000 individuals was estimated by distributing the seasonal hospitalisation rate according to weekly hospitalisation data reported in national surveillance from week 17 of 2023 to week 16 of 2024 [9]. RSV-related hospitalisation rates per 100,000 individuals in the total population and in age groups used in demographic calibration are presented in **Table S7**.

**Table S7.** Weekly hospitalisation rate, per 100,000 individuals (by total population and age group), season 2023-2024, England.

| Week | 0-4 years | 5-14 years | 15-44 years | 45-54 years | 55-64 years | 65-74 years | 75-84 years | ≥85 years | Total population |
|---|---|---|---|---|---|---|---|---|---|
| **1** | 12.472 | 0.174 | 0.234 | 0.094 | 0.835 | 1.171 | 2.858 | 10.102 | 2.678 |
| **2** | 4.663 | 0.074 | 0.286 | 0.369 | 0.923 | 1.912 | 4.719 | 8.046 | 2.170 |
| **3** | 4.264 | 0.094 | 0.196 | 0.367 | 0.820 | 1.011 | 4.468 | 11.325 | 2.022 |
| **4** | 2.435 | 0.420 | 0.065 | 0.092 | 0.719 | 1.531 | 3.434 | 7.512 | 1.540 |
| **5** | 2.051 | 0.085 | 0.033 | 0.092 | 0.102 | 1.138 | 1.064 | 5.932 | 0.869 |
| **6** | 2.860 | 0.194 | 0.067 | 0.095 | 0.000 | 0.132 | 0.223 | 1.130 | 0.517 |
| **7** | 2.635 | 0.090 | 0.242 | 0.196 | 0.109 | 0.542 | 0.000 | 0.000 | 0.641 |
| **8** | 1.386 | 0.164 | 0.063 | 0.000 | 0.000 | 0.122 | 0.205 | 2.079 | 0.366 |
| **9** | 0.640 | 0.094 | 0.033 | 0.094 | 0.209 | 0.519 | 0.871 | 1.638 | 0.437 |
| **10** | 0.640 | 0.189 | 0.065 | 0.092 | 0.000 | 0.126 | 0.638 | 0.539 | 0.295 |
| **11** | 2.051 | 0.085 | 0.065 | 0.000 | 0.000 | 0.126 | 0.213 | 1.079 | 0.379 |
| **12** | 1.373 | 0.081 | 0.000 | 0.000 | 0.099 | 0.240 | 0.403 | 2.500 | 0.385 |
| **13** | 0.677 | 0.092 | 0.000 | 0.103 | 0.000 | 0.000 | 0.000 | 1.816 | 0.198 |
| **14** | 0.657 | 0.097 | 0.000 | 0.000 | 0.105 | 0.000 | 0.000 | 1.124 | 0.164 |
| **15** | 1.052 | 0.000 | 0.067 | 0.000 | 0.000 | 0.000 | 0.660 | 0.000 | 0.230 |
| **16** | 1.085 | 0.000 | 0.000 | 0.000 | 0.108 | 0.000 | 0.000 | 0.586 | 0.165 |
| **17** | 0.455 | 0.000 | 0.000 | 0.000 | 0.000 | 0.000 | 0.000 | 0.000 | 0.049 |
| **18** | 0.417 | 0.000 | 0.000 | 0.000 | 0.000 | 0.122 | 0.000 | 0.509 | 0.090 |
| **19** | 0.981 | 0.111 | 0.000 | 0.000 | 0.000 | 0.000 | 0.000 | 0.000 | 0.139 |
| **20** | 0.638 | 0.000 | 0.000 | 0.000 | 0.000 | 0.000 | 0.000 | 0.728 | 0.095 |
| **21** | 0.366 | 0.000 | 0.000 | 0.000 | 0.000 | 0.000 | 0.000 | 0.000 | 0.040 |
| **22** | 0.000 | 0.000 | 0.000 | 0.000 | 0.000 | 0.000 | 0.000 | 0.000 | 0.184 |

| Week | 0-4 years | 5-14 years | 15-44 years | 45-54 years | 55-64 years | 65-74 years | 75-84 years | ≥85 years | Total population |
|---|---|---|---|---|---|---|---|---|---|
| **23** | 0.829 | 0.000 | 0.000 | 0.000 | 0.000 | 0.000 | 0.000 | 0.000 | 0.091 |
| **24** | 0.776 | 0.000 | 0.000 | 0.000 | 0.000 | 0.000 | 0.000 | 0.000 | 0.084 |
| **25** | 0.000 | 0.143 | 0.000 | 0.000 | 0.000 | 0.000 | 0.000 | 0.000 | 0.040 |
| **26** | 0.000 | 0.000 | 0.000 | 0.000 | 0.000 | 0.000 | 0.000 | 0.000 | 0.184 |
| **27** | 0.340 | 0.000 | 0.107 | 0.000 | 0.000 | 0.000 | 0.000 | 0.000 | 0.110 |
| **28** | 0.829 | 0.000 | 0.000 | 0.000 | 0.000 | 0.000 | 0.000 | 0.000 | 0.091 |
| **29** | 0.766 | 0.000 | 0.000 | 0.000 | 0.000 | 0.000 | 0.000 | 0.000 | 0.084 |
| **30** | 0.802 | 0.000 | 0.088 | 0.000 | 0.000 | 0.170 | 0.000 | 0.000 | 0.178 |
| **31** | 1.946 | 0.000 | 0.000 | 0.000 | 0.000 | 0.000 | 0.000 | 0.736 | 0.245 |
| **32** | 0.802 | 0.111 | 0.044 | 0.000 | 0.000 | 0.000 | 0.000 | 0.000 | 0.148 |
| **33** | 1.258 | 0.000 | 0.000 | 0.000 | 0.155 | 0.000 | 0.310 | 0.000 | 0.206 |
| **34** | 0.774 | 0.000 | 0.000 | 0.000 | 0.000 | 0.225 | 0.000 | 0.000 | 0.119 |
| **35** | 2.048 | 0.000 | 0.000 | 0.000 | 0.000 | 0.000 | 0.000 | 0.000 | 0.226 |
| **36** | 1.268 | 0.111 | 0.040 | 0.000 | 0.000 | 0.000 | 0.000 | 0.000 | 0.193 |
| **37** | 1.755 | 0.000 | 0.048 | 0.000 | 0.000 | 0.000 | 0.311 | 0.000 | 0.259 |
| **38** | 3.901 | 0.000 | 0.076 | 0.000 | 0.000 | 0.000 | 0.000 | 0.000 | 0.453 |
| **39** | 7.915 | 0.000 | 0.000 | 0.000 | 0.131 | 0.000 | 0.272 | 0.698 | 0.923 |
| **40** | 6.121 | 0.093 | 0.032 | 0.091 | 0.000 | 0.378 | 0.213 | 0.545 | 0.834 |
| **41** | 12.550 | 0.310 | 0.072 | 0.000 | 0.000 | 0.000 | 0.473 | 0.607 | 1.531 |
| **42** | 18.769 | 0.501 | 0.035 | 0.000 | 0.000 | 0.000 | 0.913 | 0.584 | 2.254 |
| **43** | 26.995 | 1.345 | 0.144 | 0.000 | 0.114 | 0.281 | 0.473 | 1.822 | 3.489 |
| **44** | 32.194 | 0.660 | 0.262 | 0.092 | 0.307 | 0.632 | 1.702 | 3.775 | 4.316 |
| **45** | 33.473 | 1.131 | 0.360 | 0.367 | 0.615 | 0.632 | 1.489 | 3.775 | 4.748 |
| **46** | 31.965 | 1.162 | 0.609 | 0.665 | 1.477 | 2.212 | 3.518 | 7.303 | 5.696 |
| **47** | 39.017 | 1.131 | 0.131 | 0.643 | 1.639 | 1.517 | 4.468 | 10.247 | 6.224 |
| **48** | 44.831 | 1.127 | 0.236 | 0.096 | 1.180 | 1.990 | 7.347 | 11.705 | 7.264 |
| **49** | 32.943 | 1.523 | 0.415 | 0.687 | 2.182 | 2.845 | 5.706 | 8.177 | 6.317 |
| **50** | 28.961 | 1.302 | 0.468 | 0.755 | 1.253 | 2.472 | 7.255 | 11.786 | 5.929 |
| **51** | 15.811 | 0.424 | 0.366 | 0.462 | 1.028 | 3.421 | 6.179 | 16.191 | 4.290 |
| **52** | 12.924 | 0.593 | 0.228 | 0.367 | 0.410 | 3.160 | 7.659 | 17.258 | 3.967 |

The initial values assumed for the proportion of ARD infections that progress to LRTD, based on available literature [12-14], are presented in **Table S8**. These values were adjusted during the epidemiological calibration process, and final calibrated values are presented in **Table S9**.

**Table S8**. Proportion of ARD infections that are LRTD – initial values used in calibration process.

| Age group | Calibration target |
|---|---|
| 0-4 years | 27.0% |
| 5-49 years | 18.2% |
| 50-64 years | 28.4% |
| 65-74 years | 41.1% |
| ≥75 years | 52.1% |

Abbreviations: ARD, acute respiratory disease; LRTD, lower respiratory tract disease.

Age specific severity inputs used in the model are presented in **Table S9**. Other epidemiological parameters are presented in **Table S10**.

**Table S9**. Key age-stratified severity inputs.

| | Parameter | Value | Source |
|---|---|---|---|
| $1-\rho$ | Proportion of ARD among RSV infections* | | |
| | 0-4 years | 84.3% | [15] |
| | 5-14 years | 47.9% | [15] |
| | 15-39 years | 23.7% | [15] |
| | 40-54 years | 22.2% | [15] |
| | 55-64 years | 29.7% | [16] |
| | 65-74 years | 44.3% | [16] |
| | ≥75 years | 76.3% | [16] |
| $\kappa$ | Proportion of LRTD among ARD infections – main analysis | | |
| | 0-4 years | 15.6% | Calibrated |
| | 5-14 years | 3.8% | Calibrated |
| | 15-44 years | 4.2% | Calibrated |
| | 45-49 years | 2.6% | Calibrated |
| | 50-54 years | 4.1% | Calibrated |
| | 55-59 years | 5.4% | Calibrated |
| | 60-64 years | 6.9% | Calibrated |
| | 65-74 years | 14.6% | Calibrated |
| | 75-84 years | 23.8% | Calibrated |
| | ≥85 years | 60.5% | Calibrated |
| $\kappa$ | Proportion of LRTD among ARD infections – exploratory analysis | | |
| | 0-4 years | 16.3% | Calibrated |
| | 5-14 years | 4.0% | Calibrated |
| | 15-44 years | 4.6% | Calibrated |
| | 45-49 years | 3.1% | Calibrated |
| | 50-54 years | 4.8% | Calibrated |
| | 55-59 years | 6.1% | Calibrated |
| | 60-64 years | 7.7% | Calibrated |
| | 65-74 years | 16.1% | Calibrated |
| | 75-84 years | 27.4% | Calibrated |
| | ≥85 years | 68.2% | Calibrated |
| $\delta$ | Proportion of hospitalisations among LRTD infections | | |
| | 0-4 years | 10.8% | [17] |
| | 5-49 years | 2.1% | [10,12,13] |
| | 50-64 years | 5.9% | [10,12,13] |
| | 65-74 years | 10.1% | [10,12,13] |
| | ≥75 years | 18.7% | [10,12,13] |
| $\mu$ | Proportion of deaths among hospitalized cases | | |
| | 0-19 years | 0.1% | [18] |
| | 20-49 years | 25.0% | [12,13] |
| | 50-64 years | 20.0% | [12,13] |
| | 65-74 years | 33.7% | [12,13] |
| | ≥75 years | 66.2% | [12,13] |

**Abbreviations**: ARD, acute respiratory disease; LRTD, lower respiratory tract disease; RSV, respiratory syncytial virus.

* As the available sources reported the proportion of asymptomatic RSV infections, the proportion of ARD among all RSV infections was estimated as the complement, by subtracting the proportion of asymptomatic cases from 100%.

**Table S10**. Key epidemiological parameters not stratified by age.

| | Parameter | Value | Source |
|---|---|---|---|
| $1/\eta_m$ | Duration of maternal immunity (days) | 97 | [19] |
| $1/\varepsilon$ | Exposure to infection (Duration of latency period, days) | 4.0035 | [19] |
| $\sigma$ | Relative risk of infection following previous, infections | | |
| | 1st infection | 0.76 | [20-23] |
| | 2nd infection | 0.60 | |
| | 3rd + infection | 0.40 | |
| $1/\eta_m$ | Duration of infection (days) | | |
| | 1st infection | 7.54 | [19] |
| | 2nd infection | 6.52 | [19] |
| | 3rd + infection | 4.04 | [19] |
| $1/\eta$ | Duration of antibody protection for those previously infected (days) | 130 | [19] |
| $\psi$ | Relative infectiousness | | |
| | 2nd infection | 0.75 | [24] |
| | 3rd + infection | 0.51 | [24] |
| $b$ | Transmission probability – main analysis | 0.08 | Calibrated |
| | Transmission probability – exploratory analysis (demographic shift) | 0.08 | Calibrated |
| $s$ | Seasonality parameters | | |
| | Amplitude – main analysis | 0.26 | Calibrated |
| | Amplitude – exploratory analysis (demographic shift) | 0.25 | Calibrated |
| | Shift– main analysis | 0.75 | Calibrated |
| | Shift – exploratory analysis (demographic shift) | 0.74 | Calibrated |

##### *S1.3 Model calibration*

##### *S1.4 Calibration of demographic parameters*

All-cause mortality and migration in the UK was calibrated to reproduce the population structure. Calibration was informed by the data on population size by age group, for a starting year (1994) and a target year (2023 for the main analysis, 2030 for the exploratory analysis). For the demographic calibration process, the dynamic transmission model (DTM) was initialized with population size and distribution for 1994, within compartments $M$ and $S_1$: it was assumed that 10% of age group 0-4 years is in $M$ compartment and 90% of age group 0-4 years and all other age groups are in $S_1$ compartment. RSV infection was not included for demographic calibration.

The rate of population change by age group was calibrated to obtain age structure similar to that observed in a target year, using R function *optim*. The initial parameter values for optimization were defined in the code by the user. The number of individuals in each age group in a target year, as simulated by the model, was compared to the actual data (reported or projected for a target year), and the goodness of model fit was assessed using a likelihood function. The R function *optim* algorithm estimated new initial values for the next iteration and repeated the process until the maximum number of iterations was reached or a satisfactory result was achieved, as defined via maximization of a likelihood function.

Calibration of demographic parameters was conducted for the main analysis, and for an exploratory analysis on demographic shift.

##### *S1.5 Calibration of epidemiological parameters*

Three transmission parameters were calibrated to reproduce RSV incidence in population over time: (1) transmission probability from an infectious to a susceptible individual, (2) the amplitude of the seasonal cosine function to capture variations in infection intensity, and (3) the horizontal shift of the seasonal cosine function used to define infection seasonality at each week of the simulation. Calibration was performed using R function *optim*, in line with methodology described by Pitzer et al., [24]

The calibration target was set to the expected number of RSV-related hospitalisations, derived from Osei-Yeboah et al. [11], a reported hospitalisation rate [9] underreporting rate [10] and population size for each year of a calibration timeframe. In epidemiological calibration process the DTM was initialized by introducing a single infectious person, in a compartment $I_1$, in each age group. The RSV transmission was then simulated over the model time horizon.

Calibration timeframe, i.e., a time period used to compare simulated number of hospitalisations to the expected number of hospitalisations, was set to a 10-year period of 2023-2032.

Initial values for transmission probability, seasonal amplitude and seasonal shift were set to 0.1, 0.2, and 0.7, respectively.

In the first iteration, the model was run with the defined initial values over the period of 1994-2032. Then, a number of hospitalisations in each week in the calibration timeframe, as simulated by the model, was compared with the expected number of hospitalisations for 2023-2032, and the goodness of model fit was assessed using a likelihood function. The algorithm of R function *optim* estimated new initial values for the next iteration and repeated the process until the maximum number of iterations was reached or a satisfactory result was achieved, as defined via maximization of a likelihood function.

Calibration of epidemiologic parameters was performed using a step-wise procedure, guided by two main criteria: (1) fit of the simulated number of hospitalisations in the total population to the expected number of hospitalisations; (2) simulated age distribution of hospitalisations deviated from the expected age distribution. In case if the criterion on age distribution was not met, proportion of ARD infections that are LRTD was adjusted by a factor accounting for difference between simulated and expected number of hospitalisations, computed by age group. This model input was chosen for adjustment, as the previous literature review suggested that it is highly uncertain due to limited data availability. After this adjustment, model calibration was re-run, until a difference between simulated and actual number of hospitalisations in total population and in age group was acceptable. For this analysis, a ~5% deviation was considered as a target threshold.

Calibration of epidemiological parameters was conducted for the main analysis, and for an exploratory analysis on demographic shift.

##### *S1.6 Modelling of vaccination*

The model accounted for three types of vaccine protection specific to disease severity:

- Protection against RSV infection (symptomatic ARD or asymptomatic) – implemented as a relative risk of infection in vaccinated versus unvaccinated individuals, by time since vaccination ($\sigma_4^j$, where j is the number of weeks since vaccination);
- Protection against LRTD infection given ARD – implemented as a relative risk of LRTD given ARD, in vaccinated versus unvaccinated individuals, by time since vaccination ($\sigma_5^j$, where j is the number of weeks since vaccination);
- Protection against hospitalisation given LRTD – implemented as a relative risk of hospitalisation given LRTD, in vaccinated versus unvaccinated individuals, by time since vaccination ($\sigma_6^j$, where j is the number of weeks since vaccination).

Waning of post-vaccination immunity was explicitly modelled for each type of vaccine protection. The model allows consideration of weekly changes of the level of protection over 3 years (156 weeks) since vaccination, and a stable protection for the following period (157+ weeks).

Vaccine efficacy inputs were informed by a pivotal Phase 2/3 ConquerRSV trial for mRNA-1345, a case-driven, randomized, double-blind, placebo-controlled, multi-continent study, aiming to evaluate the safety and efficacy of the vaccine as compared with placebo in adults ≥ 60 years of age [25,26].

###### S1.6.1 Vaccine efficacy, main analysis

In the main analysis, a non-linear waning model was adopted to represent the time-dependent decline in vaccine efficacy. This approach is supported by accumulating evidence from multiple Phase 2/3 mRNA-1345 clinical studies, which indicate that protection provided by the vaccine does not decline at a constant rate. To accurately reflect this biologically plausible behavior, a non-linear function was incorporated into the model:

$$VE(t) = 1 - \exp\left(\beta_0 + \beta_1 \cdot \log(t)\right)$$

where $VE(t)$ is vaccine efficacy at time t in days, $\beta_0$ represents the initial log-odds of protection, $\beta_1$ captures the rate of decline in efficacy over time on the logarithmic scale.

Parameters of this function are provided in **Table S11**.

Estimated vaccine protection by week is presented in **Figure S2**.

**Table S11**. Vaccine efficacy inputs – non-linear wanning

| Parameter | Against ARD | Against LRTD |
|---|---|---|
| $\beta_0$ | -1.9787039 | -2.5107449 |
| $\beta_1$ | 0.234412 | 0.3057026 |

**Abbreviations**: ARD, acute respiratory disease; LRTD, lower respiratory tract disease.

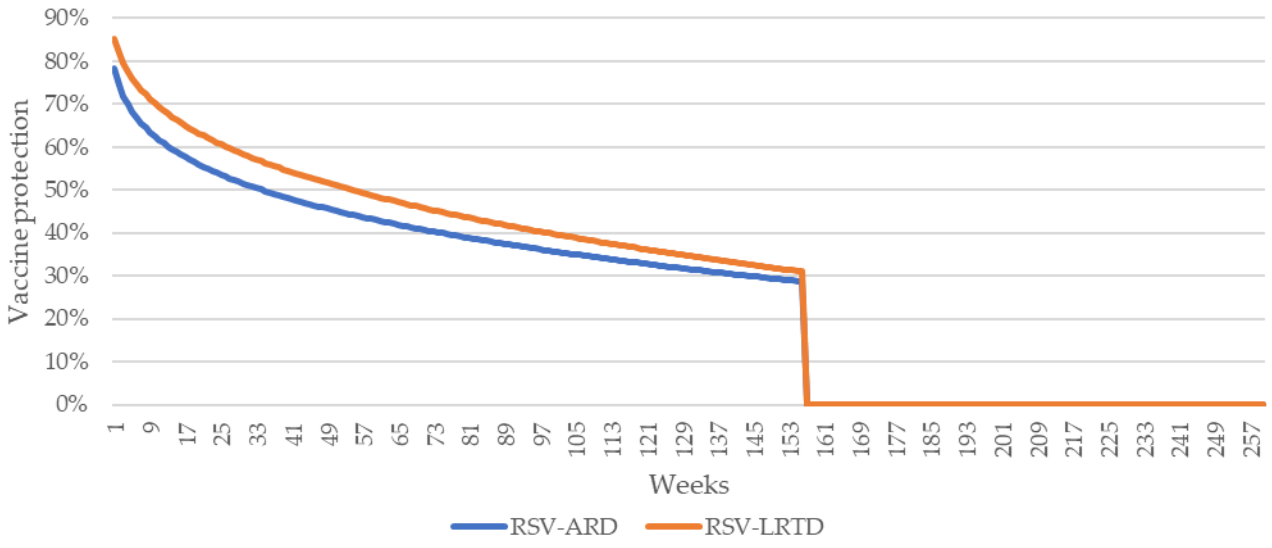


**Figure S2**. Vaccine protection by week since vaccination – main analysis

S1.6.2 Vaccine efficacy, exploratory analysis (alternative VE)

In the exploratory analysis, an alternative assumption on vaccine efficacy was tested, which included a third type of efficacy (against hospitalizations) and linear waning for all types of protection.

The initial vaccine efficacy at month 0 was set equal to the efficacy reported in the primary analysis of the ConquerRSV trial. Vaccine efficacy against RSV-LRTD with ≥2 symptoms was assessed at regular intervals over the follow-up period. A weighted least squares regression was performed on the vaccine efficacy estimates, using inverse variance weighting based on placebo case data. This approach produced monthly waning rates of 1.9% for RSV-LRTD and RSV hospitalisation, and 1.8% for RSV-ARD.

This waning rate was applied to linear decline of vaccine protection for the more severe endpoint of RSV-LRTD requiring inpatient care and the less severe endpoint of RSV-ARD.

Further, assumptions on initial vaccine protection and waning of protection were applied to derive inputs for the DTM, which required weekly estimates of vaccine efficacy, starting from week 1 post-vaccination. A linear waning function constructed as described above was used to compute vaccine efficacy function dependent on the number of weeks after vaccination.

Estimated vaccine protection by week is presented in **Figure S3**.

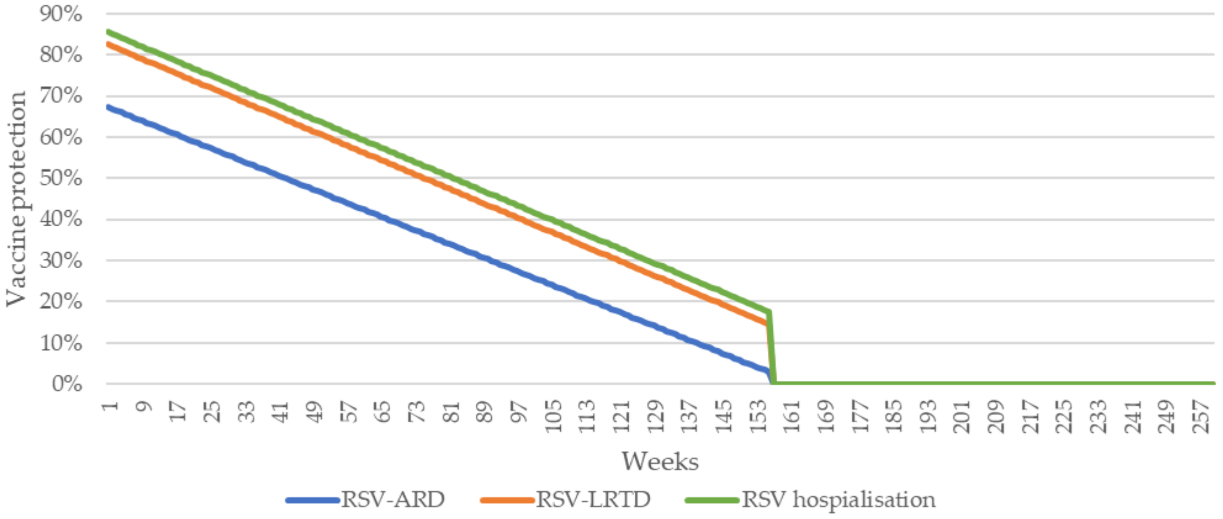


**Figure S3**. Vaccine protection by week since vaccination – exploratory analysis

S1.6.3 Vaccination coverage

An RSV vaccination coverage of 60% was assumed for individuals aged 65 years and older, consistent with observed RSV coverage measured as of 31 March 2025 [27]. Additionally, an alternative scenario tested uptake levels aligned with influenza vaccine coverage during the 2024-2025 season, estimated at approximately 80% [28].

To inform the weekly progression of RSV vaccination coverage in the model, cumulative influenza vaccination uptake data for individuals aged 65 years and older were used [29]. The number of influenza vaccine doses administered each week, from the start of the 2024-2025 influenza campaign through week 17, was expressed as a proportion of the total cumulative influenza coverage. These weekly proportions were then scaled to the 60% or 80% coverage target, under the assumption that RSV vaccination would follow a similar temporal pattern. Weekly RSV vaccination coverage inputs and corresponding influenza uptake data are presented in **Table S12**.

**Table S12**. Inputs for RSV vaccination coverage based on weekly influenza uptake in the 2024-2025 season.

| Week since a start of vaccination programme | Cumulative number of influenza vaccinations administered | Proportion of final influenza vaccine uptake* | Estimated cumulative RSV vaccine coverage – flu uptake | Estimated cumulative RSV vaccine coverage – influenza uptake |
|---|---|---|---|---|
| 1 | 3,931,614 | 45.6% | 27.4% | 36.5% |
| 2 | 5,453,986 | 63.3% | 38.0% | 50.7% |
| 3 | 6,363,851 | 73.9% | 44.3% | 59.1% |
| 4 | 7,374,237 | 85.6% | 51.4% | 68.5% |
| 5 | 7,764,619 | 90.1% | 54.1% | 72.1% |
| 6 | 8,019,298 | 93.1% | 55.9% | 74.5% |
| 7 | 8,190,996 | 95.1% | 57.1% | 76.1% |
| 8 | 8,316,351 | 96.5% | 57.9% | 77.2% |
| 9 | 8,396,674 | 97.5% | 58.5% | 78.0% |
| 10 | 8,480,678 | 98.5% | 59.1% | 78.8% |
| 11 | NA | NA | 59.3% | 79.0%** |
| 12 | NA | NA | 59.4% | 79.3%** |
| 13 | 8,559,430 | 99.4% | 59.6% | 79.5% |
| 14 | 8,583,279 | 99.6% | 59.8% | 79.7% |
| 15 | 8,598,081 | 99.8% | 59.9% | 79.9% |
| 16 | 8,608,673 | 99.9% | 60.0% | 80.0% |
| 17 | 8,613,876 | 100.0% | 60.0% | 80.0% |

**Abbreviations**: NA, data not available for weeks 11 and 12; RSV, respiratory syncytial virus.

*Estimated assuming that the cumulative number of influenza vaccinations administered by week 17 represents 100% of total uptake.

**Estimated by linear interpolation between weeks 10 and 13.

### S2 Supplement 2: Additional model results

#### *S2.1 Model fit and calibration outputs*

The model demonstrated a good fit to demographic and epidemiologic data considered for calibration.

Figures below (**Figure S4** to **Figure S7)** present results of demographic calibration. For the total population size, differences between the actual and simulated estimates ranged from 1% to 8% in the main analysis (population size slightly underestimated), and from −3% to 2% in the exploratory analysis on demographic shift (population size slightly overestimated for 2024 and underestimated for 2044). Simulated population size by age group closely corresponded to the target age distribution (as reported in 2023 for the main analysis or projected in 2030 for the exploratory analysis).

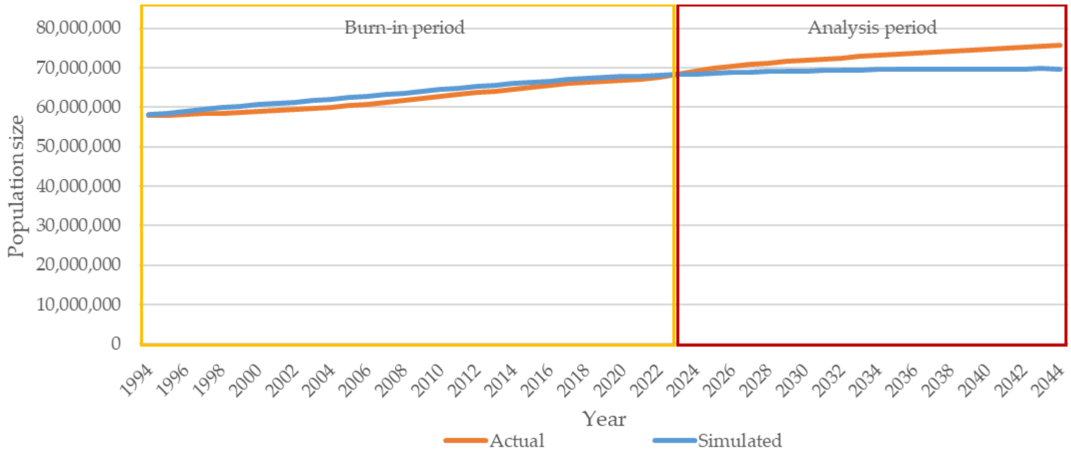


**Figure S4**. Total population size: comparison of model projections and target values over time (1994–2044) – main analysis.
**Note:** Actual data represent the reported population size, for1994-2023 and projected population size for 2024-2044.

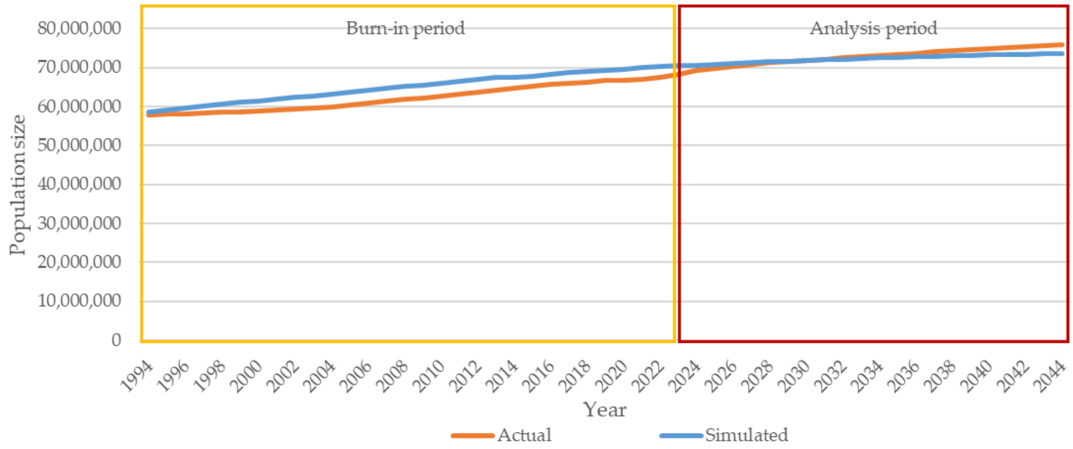


**Figure S5**. Total population size: comparison of model projections and target values over time (1994-2044) – exploratory analysis on demographic shift.
**Note:** Actual data represent the reported population size for 1994-2023, and projected population size for 2024-2044.

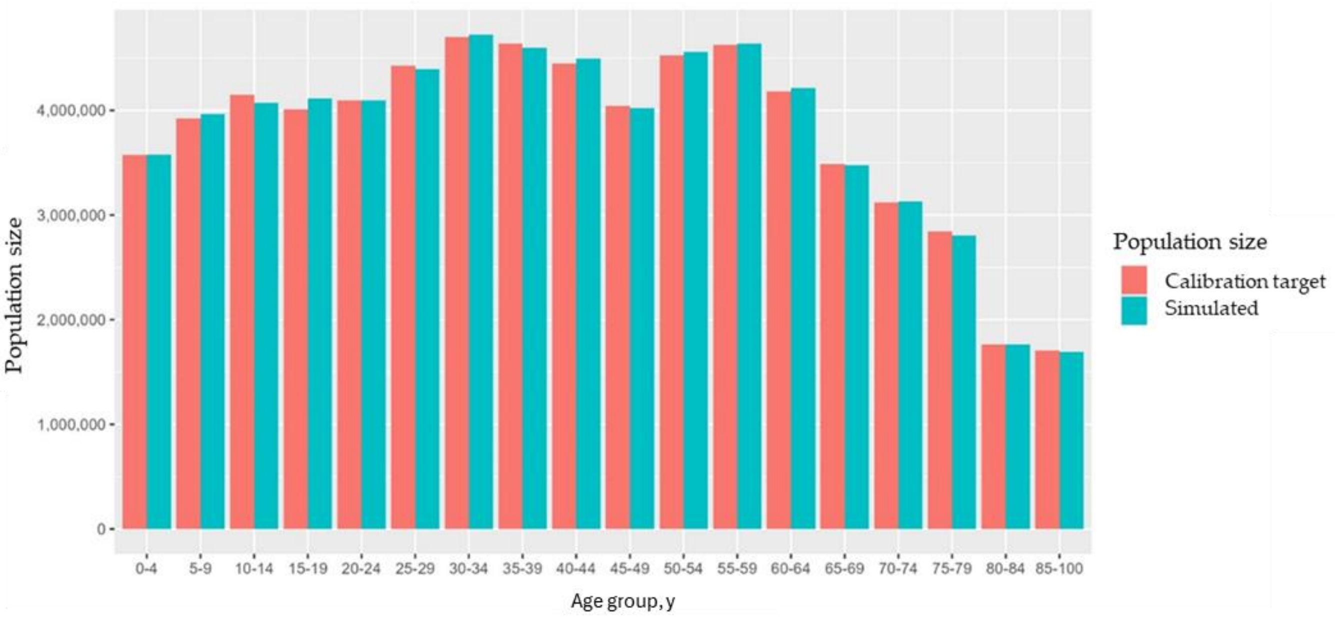


**Figure S6**. Population size by age group: comparison of model projections and calibration target values in 2023 – main analysis. **Abbreviations:** y, years.

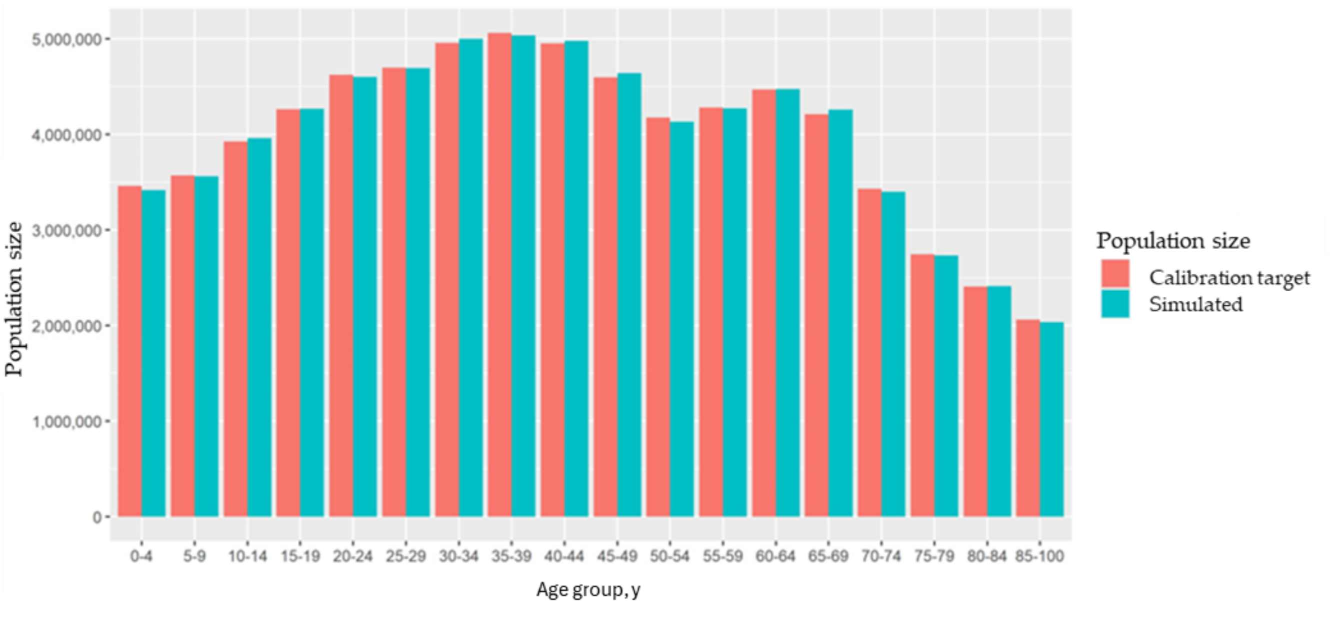


**Figure S7**. Population size by age group: comparison of model-simulated and calibration target values in 2030 – exploratory analysis on demographic shift.

**Abbreviations:** y, years.

Prior to the start of the analytic time horizon, the model was run over a 30-year burn-in period (staring 1994), to allow for stabilization of RSV transmission dynamics. As shown in **Figure S8**, the designated analysis period corresponds to the phase in which stabilisation of the key epidemiological indicators was achieved.

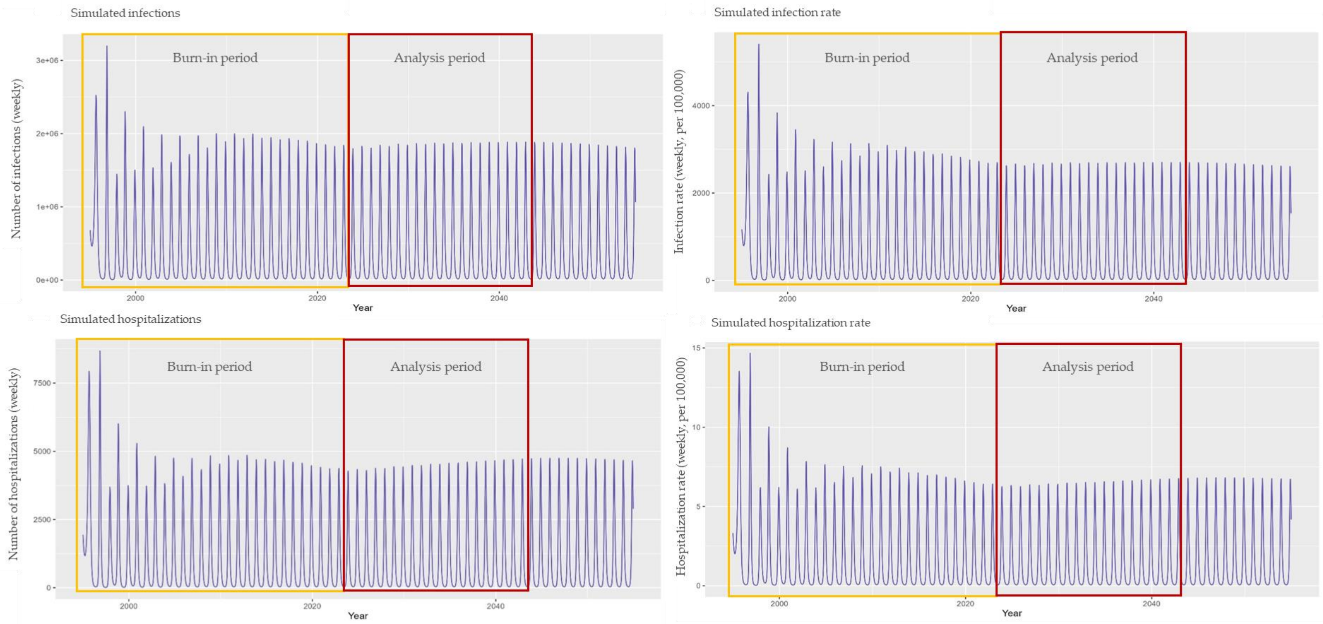


**Figure S8.** Simulated incidence of RSV infections and hospitalizations over the full model timeframe.

Figures below (**Figure S9** and **Figure S10**), **Table S13** and **Table S14**, present the results of epidemiologic calibration, with comparison between model projections and target hospitalization rates per 100,000 population, over the 2023–2032 calibration period. The model was calibrated to reproduce weekly hospitalisation rates in the total population, ensuring alignment with the observed seasonal patterns and allowing for the interannual variability. As shown in **Figure S9**, simulated weekly rates align closely with observed seasonal trends over the calibration period (2023–2032).

Total number of hospitalisations simulated by the model over 10 years was close to the expected actual number of hospitalisations, with a difference of 4%, which was considered acceptable (**Table S13).** Over the calibration timeframe, slight interannual variability was observed, with difference between the actual and simulated number of hospitalisations ranging from 2% to 6% across individual years.

Additionally, model inputs were refined to match age-specific hospitalisation patterns. The resulting average annual hospitalisation rates by age group, presented in **Figure S10** and **Table S14**, demonstrate strong concordance with target values.

These results support the reliability of the model's predictions of both the age distribution and temporal dynamics of RSV-related hospitalizations during the calibration period, providing a robust foundation for projections over the full analysis horizon, spanning from the 2024-2025 to 2043-2044 RSV seasons.

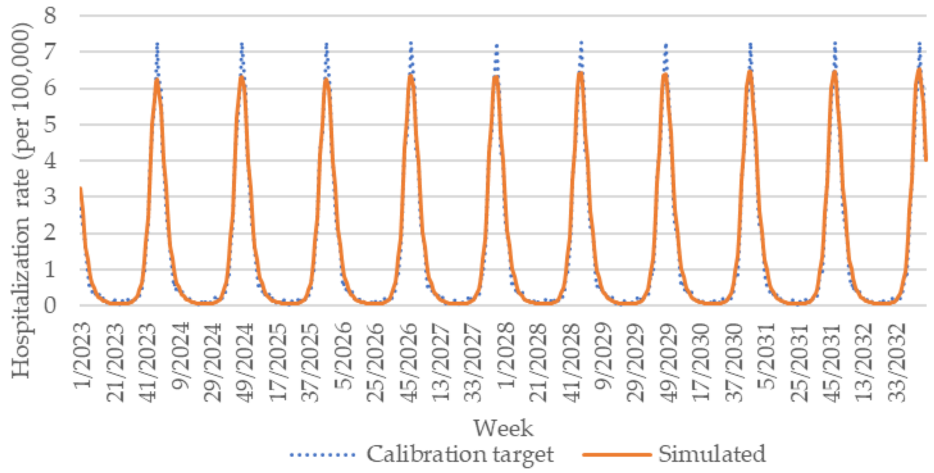


**Figure S9.** Weekly hospitalization rates per 100,000 population: comparison of model projections and calibration target values over time (2023–2032).

**Table S13**. Number of hospitalizations: comparison of model projections and calibration target values over time (2023–2032).

| | **Overall** | **Year** | | | | | | | | | |
|---|---|---|---|---|---|---|---|---|---|---|---|
| | **2023-2032** | **2023** | **2024** | **2025** | **2026** | **2027** | **2028** | **2029** | **2030** | **2031** | **2032** |
| Simulated | **531,390** | 51,628 | 52,812 | 51,872 | 53,227 | 52,585 | 53,708 | 53,213 | 54,113 | 53,752 | 54,479 |
| Actual | **511,120** | 50,693 | 50,803 | 50,906 | 51,004 | 51,096 | 51,181 | 51,259 | 51,331 | 51,395 | 51,452 |
| Difference | **4%** | 2% | 4% | 2% | 4% | 3% | 5% | 4% | 5% | 5% | 6% |

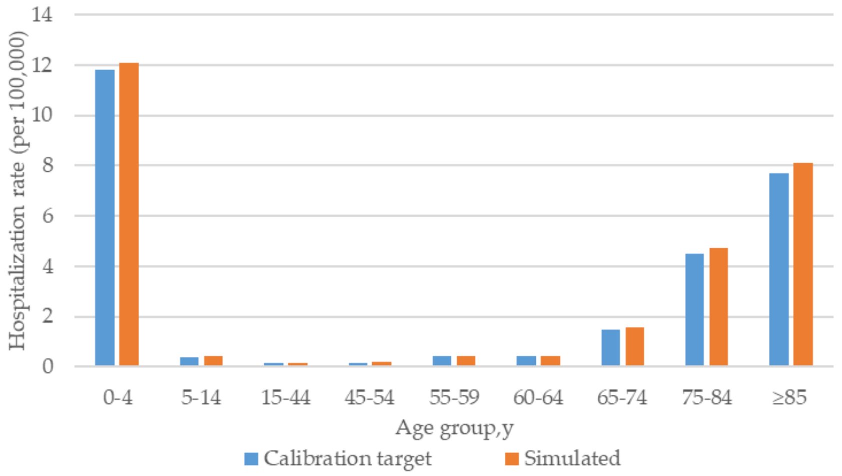


**Figure S10.** Comparison of model projections and calibration target hospitalization rates by age group (per 100,000 population), based on the average annual rate over the 2023–2032 calibration period.
**Abbreviations**: y, years.

**Table S14**. Total number of hospitalisations (2023–2032): comparison of model projections and calibration target values by age group

| | Overall | Age group, years | | | | | | | | |
|---|---|---|---|---|---|---|---|---|---|---|
| | **2023-2032** | **0-4** | **5-14** | **15-44** | **45-54** | **55-59** | **60-64** | **65-74** | **75-84** | **≥85** |
| Simulated | **531,390** | 214,862 | 16,509 | 21,708 | 7,974 | 10,844 | 9,929 | 55,914 | 118,212 | 75,439 |
| Actual | **511,120** | 209,522 | 16,094 | 20,890 | 7,627 | 10,352 | 9,475 | 53,228 | 112,292 | 71,639 |
| Difference | **4%** | 3% | 3% | 4% | 5% | 5% | 5% | 5% | 5% | 5% |

**Abbreviations**: y, years.

*S2.2 Additional validation results*

Comparison of model projections and estimates reported in the literature for age-specific annual incidence rates of ARD per 100,000 individuals is presented in **Table S15**. Published values vary substantially across age groups, reflecting heterogeneity in the available data. Previous studies have highlighted inconsistencies in case definitions, data collection methods, and population sampling, which contribute to the observed variability in reported incidence rates. Despite these differences, the ARD incidence rates predicted by the DTM fall within or remain comparable to the reported ranges, indicating broad consistency between model outputs and existing evidence.

**Table S15**. ARD rate per 100,000 in model prediction and available sources.

| Age group | DTM outputs | Literature | Source |
|---|---|---|---|
| 16+ | 7,419 | 1,600-7,400 | Wilkinson 2023 et al. [30] |
| 18+ | 7,321 | 900-13,100 | Wilkinson 2023 et al. [30] |
| 18+ | 7,321 | 6,481 | RAND [31] |
| 15-24 | 9,295 | 10,300 | Wilkinson 2023 et al. [30] |
| 15-44 | 9,186 | ~2,000-6,010 | Wilkinson 2023 et al. [30] |
| 25-44 | 9,137 | 10,200 | Wilkinson 2023 et al. [30] |
| 18-49 | 8,792 | 6,252 | RAND [31] |
| 45+ | 5,970 | 6,120-7,620 | Wilkinson 2023 et al. [30] |
| 45-64 | 6,465 | 17,900 | Wilkinson 2023 et al. [30] |
| 50-64 | 6,198 | 7,073 | RAND [31] |
| 60+ | 5,385 | 4,570-5,750 | Wilkinson 2023 et al. [30] |
| 65-74 | 5,536 | 7,217 | RAND [31] |
| 75+ | 5,059 | 6,208 | RAND [31] |

**Abbreviation:** ARD, acute respiratory disease; DTM, dynamic transmission model.

**Note:** DTM outputs are presented for a season 2024/2025, for a strategy without vaccination.

ARD rate based on RAND was estimated dividing reported number of cases by average population size across the 2024/2025 season.

Comparison of model projections and estimates reported in the literature for age-specific annual hospitalisation rates per 100,000 individuals, is presented in **Table S16**. Despite variability across data sources, the hospitalisation rates predicted by the DTM were within the reported ranges, for the majority of age groups, indicating broad consistency between model outputs and available empirical evidence. Description of validation presented in the main text is focused on key independent sources. **Table S16** also includes additionally the data from United Kingdom Health Security Agency (UKHSA) upscaled by 1.5 and Osei-Yeboah et al., which were used as a calibration target, and therefore present a limited value for the model validation [32].

**Table S16**. Hospitalisation rate per 100,000 in model prediction and available sources.

| Age group, years | DTM outputs | UKHSA data [9] | UKHSA data [9], upscaled by 1.5 [10] | Osei-Yeboah et al. [11] | Howa et al. [33] | RAND [31] | Fleming et al. [12] | Sharp et al. [34] | Hodgson et al. [35] | Johannesen et al. [32] | Zhang et al. [36] |
|---|---|---|---|---|---|---|---|---|---|---|---|
| 0-4 | 608 | 406 | 610 | NA | NA | NA | NA | NA | NA | NA | NA |
| 5-9 | 23 | 14 | 21 | NA | NA | NA | NA | NA | 4 | NA | NA |
| 10-14 | 18 | | | NA | NA | NA | NA | NA | | NA | NA |
| 15-19 | 9 | 5 | 8 | NA | NA | NA | NA | NA | 30 | NA | NA |
| 20-24 | 7 | | | 7* | 8* | 4* | 3-5 | NA | | 10 | NA |
| 25-29 | 7 | | | | | | | NA | | | NA |
| 30-34 | 8 | | | | | | | NA | | | NA |
| 35-39 | 9 | | | | | | | NA | | | NA |
| 40-44 | 8 | | | | | | | NA | | | NA |
| 45-49 | 4 | 6 | 9 | | | | | NA | | | NA |
| 50-54 | 13 | | | | 32 | 30 | 22-36 | NA | | | NA |
| 55-59 | 22 | 15 | 22 | | | | | NA | | | NA |
| 60-64 | | | | | | | | NA | | | 105 |
| 65-69 | 82 | 29 | 43 | 77 | 91 | 86 | 62-101 | 71 | 71 | 90 | 128 |
| 70-74 | | | | | | | | | | | 185 |
| 75-79 | 237 | 68 | 102 | 231 | | 234 | 180-291 | 251 | 251 | 280 | 227 |
| 80-84 | | | | | | | | | | | |
| 85-100 | 405 | 152 | 228 | 395 | | | | | | 600 | 752 |
| **≥ 65** | **178** | **59** | **88** | **174** | **91** | **158** | **119-193** | **159** | **159** | **225** | **276** |
| **Total** | **75** | **32** | **47** | **NA** | **NA** | **NA** | **NA** | **NA** | **NA** | **NA** | **NA** |

**Abbreviations:** DTM, dynamic transmission model; NA, not available; UKHSA, UK Health Security Agency; y, years.

**Note:** DTM outputs are presented for the 2024-2025, for a strategy without vaccination.

Hospitalisation rate for individuals aged 65 years and older was estimated using data reported in the literature and simulated population size in this age group at the end of 2024-2025 season (week 9 of 2025).

*For the RAND, Howa et al., and Osei-Yeboah et al., hospitalisation rates were originally reported for populations aged 18 years and older. In this analysis, values are presented starting from 20 years of age to match the age stratification used in the model.

##### *S2.3 RSV vaccination strategies: impact in targeted and total population*

The number of cases, number of cases avoided, and percentage of cases avoided in the target and total population with a vaccine coverage >80% are presented in **Table S17** and **Table S18,** respectively.

**Table S17.** Number of cases, number of cases avoided, and percentage of cases avoided in the **target population**, mRNA-1345 vs no vaccination over 20 years

| **Strategy** | **No vaccination** | | **Strategy 5** | **Strategy 6** | **Strategy 7** | **Strategy 8** |
|---|---|---|---|---|---|---|
| **Age group** | **75-80 y** | **≥ 60 y** | **≥ 60 y** | **75-80 y** | **75-80 y** | **≥ 60 y** |
| **Coverage** | **-** | **-** | **60%** | **80%** | **60%** | **80%** |
| **Revaccination** | **-** | **-** | **No** | **No** | **Every 3 y** | **Every 3 y** |
| **Number of cases** | | | | | | |
| RSV ARD | 4,195,188 | 20,053,473 | 18,358,175 | 3,450,965 | 3,164,318 | 12,499,634 |
| RSV LRTD | 997,987 | 3,725,497 | 3,444,496 | 800,336 | 725,173 | 2,113,360 |
| RSV hospitalisation | 186,227 | 552,758 | 514,920 | 149,345 | 135,319 | 307,534 |
| RSV death | 123,356 | 318,698 | 298,031 | 98,925 | 89,635 | 175,004 |

| | | | | | | |
|---|---|---|---|---|---|---|
| **Number of cases avoided** | | | | | | |
| RSV ARD | - | - | 1,695,298 | 744,224 | 1,030,870 | 7,553,838 |
| RSV LRTD | - | - | 281,001 | 197,651 | 272,814 | 1,612,137 |
| RSV hospitalisation | - | - | 37,838 | 36,882 | 50,908 | 245,225 |
| RSV death | - | - | 20,667 | 24,431 | 33,721 | 143,694 |
| **Percentage of cases avoided** | | | | | | |
| RSV ARD | - | - | 8% | 18% | 25% | 38% |
| RSV LRTD | - | - | 8% | 20% | 27% | 43% |
| RSV hospitalisation | - | - | 7% | 20% | 27% | 44% |
| RSV death | - | - | 6% | 20% | 27% | 45% |

**Abbreviations**: ARD, acute respiratory disease; LRTD, lower respiratory tract disease; y, years; RSV, respiratory syncytial virus; y, years.

**Note**: Percentages of cases avoided are calculated using the total number of cases without vaccination as a denominator, for each respective target age group. Due to variation in population size, percentages are not directly comparable between strategies with different eligibility criteria.

**Table S18.** Number of cases, number of cases avoided, and percentage of cases avoided in the **total population**, mRNA-1345 vs no vaccination, over 20 years

| **Strategy** | **No vaccination** | **Strategy 5** | **Strategy 6** | **Strategy 7** | **Strategy 8** |
|---|---|---|---|---|---|
| **Age group** | **Total population** | **≥ 60 y** | **75-80 y** | **75-80 y** | **≥ 60 y** |
| **Coverage** | **-** | **60%** | **80%** | **60%** | **80%** |
| **Revaccination** | **-** | **No** | **No** | **Every 3 y** | **Every 3 y** |
| **Number of cases** | | | | | |
| RSV ARD | 153,877,864 | 151,034,407 | 152,558,805 | 151,307,205 | 142,031,599 |
| RSV LRTD | 12,025,486 | 11,681,296 | 11,752,142 | 11,430,074 | 10,178,790 |
| RSV hospitalisation | 1,090,248 | 1,048,844 | 1,042,852 | 986,686 | 831,911 |
| RSV death | 336,077 | 315,171 | 305,694 | 269,598 | 191,499 |
| **Number of cases avoided** | | | | | |
| RSV ARD | - | 2,843,456 | 1,319,059 | 2,570,659 | 11,846,264 |
| RSV LRTD | - | 344,189 | 273,344 | 595,411 | 1,846,696 |
| RSV hospitalisation | - | 41,403 | 47,396 | 103,562 | 258,336 |
| RSV death | - | 20,906 | 30,384 | 66,479 | 144,578 |
| **Percentage of cases avoided** | | | | | |
| RSV ARD | - | 2% | 1% | 2% | 8% |
| RSV LRTD | - | 3% | 2% | 5% | 15% |
| RSV hospitalisation | - | 4% | 4% | 9% | 24% |
| RSV death | - | 6% | 9% | 20% | 43% |

**Abbreviations**: ARD, acute respiratory disease; LRTD, lower respiratory tract disease; RSV, respiratory syncytial virus; y, years.

**Figure S11.** Percentage of cases avoided in **total population**, mRNA-1345 vs no vaccination (over 20 years).

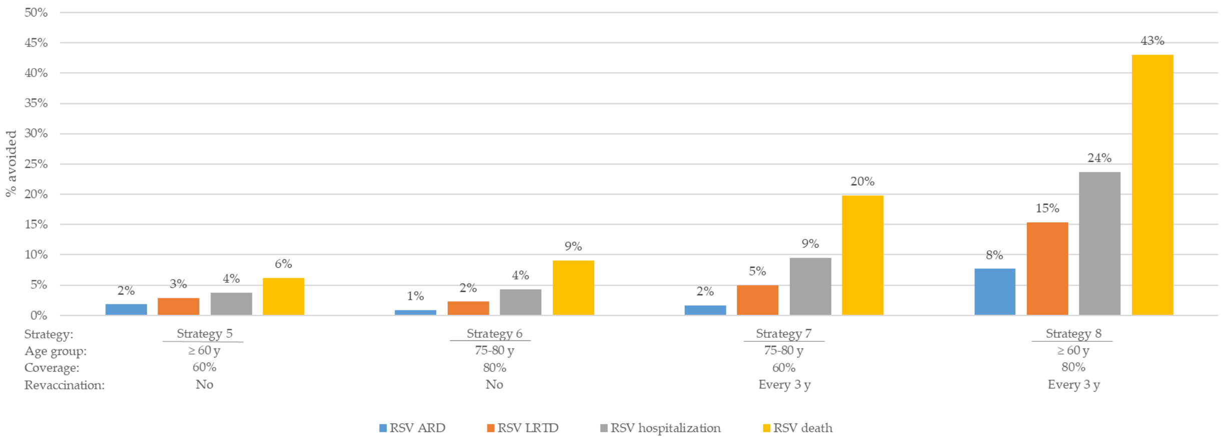


**Abbreviations:** ARD, Acute respiratory disease; LRTD, Lower respiratory tract disease; RSV, Respiratory syncytial virus; y, years.

**Figure S12.** Percentage of cases avoided in **total population** by age group, mRNA-1345 vs no vaccination (over 20 years).

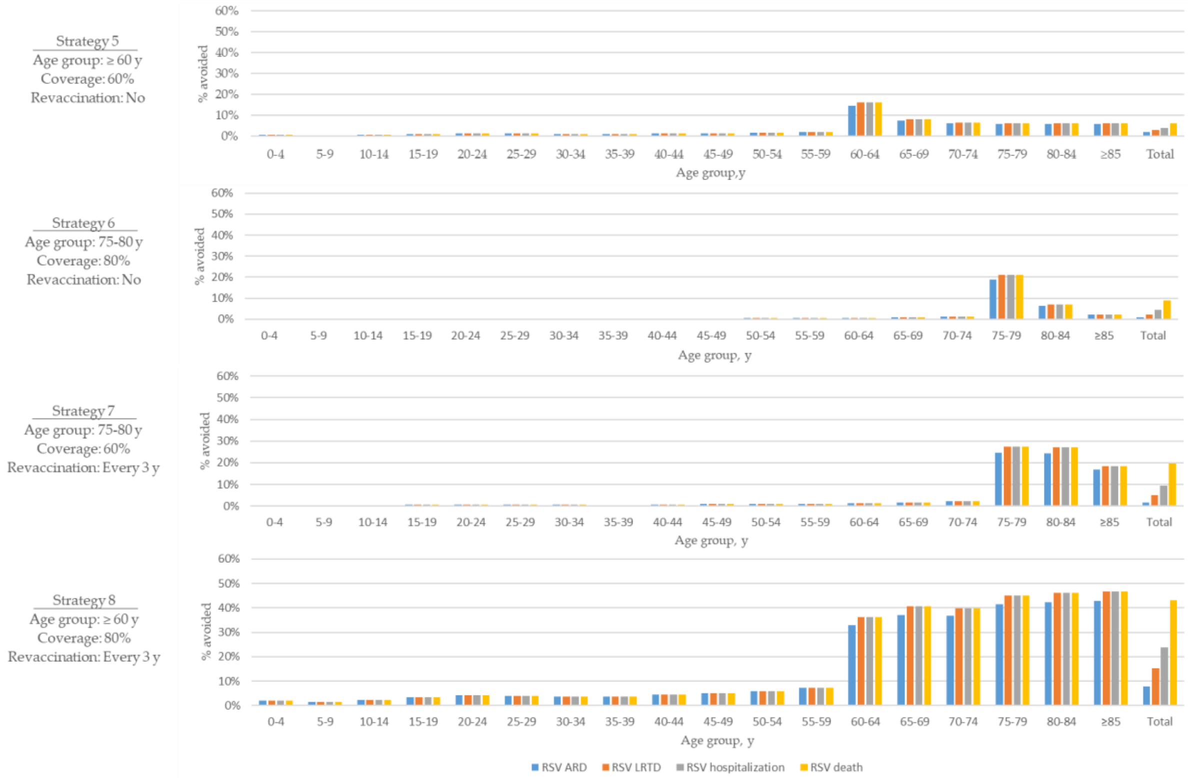


**Abbreviations**: ARD, Acute respiratory disease; LRTD, Lower respiratory tract disease; RSV, Respiratory syncytial virus; y, years.

**Figure S13**. Percentage of cases avoided in **total population** over time, mRNA-1345 vs no vaccination.

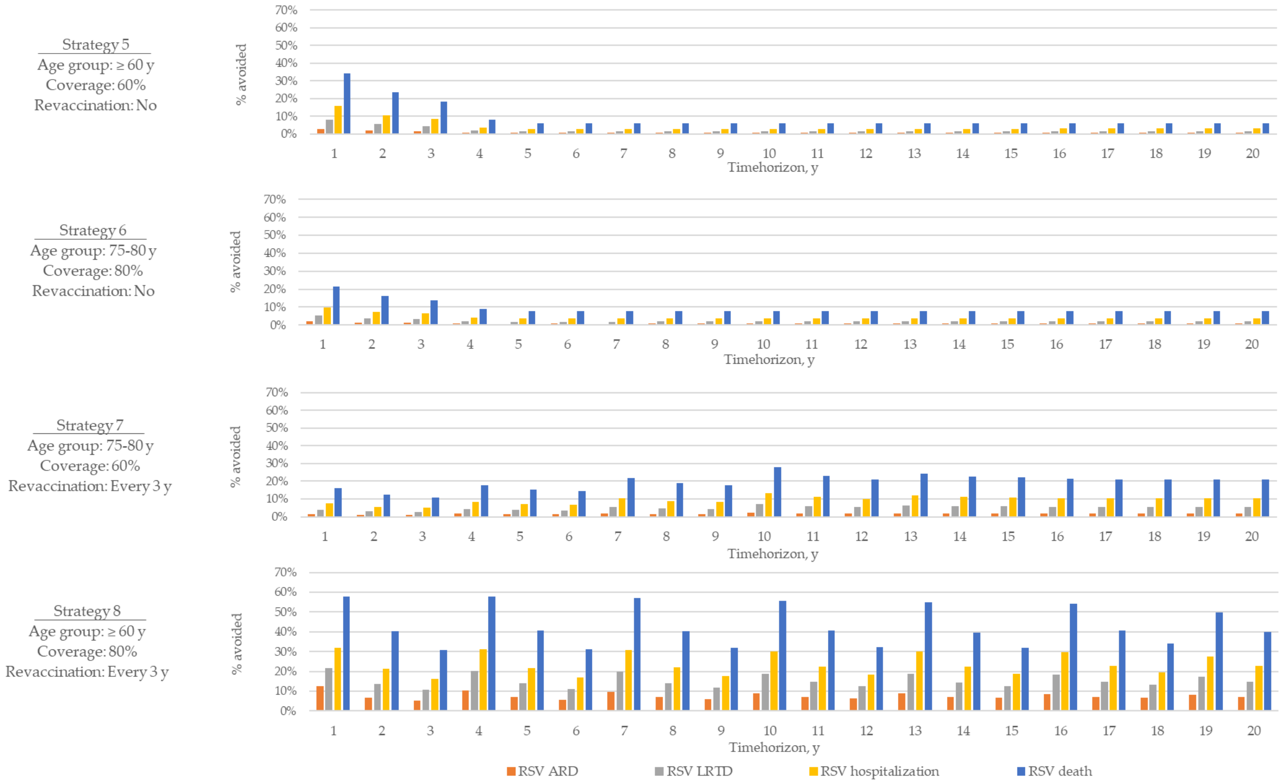


**Abbreviations**: ARD, Acute respiratory disease; LRTD, Lower respiratory tract disease; RSV, Respiratory syncytial virus; y, years.

##### *S2.4 Number of vaccine doses administered*

The total number of vaccine doses administered under each strategy over a 20-year period is presented in **Table S19**. Dose counts varied substantially across strategies, from approximately 9 million with the most restrictive approach (Strategy 1) to over 150 million under the strategy with revaccination every 2 years (Strategy 4).

**Table S19**. Number of vaccine doses in total population over 20 years.

| **Strategy** | **Strategy 1** | **Strategy 2** | **Strategy 3** | **Strategy 4** | **Strategy 5** | **Strategy 6** | **Strategy 7** | **Strategy 8** |
|---|---|---|---|---|---|---|---|---|
| **Age group** | **75-80 y** | **≥ 75 y** | **75-80 y** | **≥ 60 y** | **≥ 60 y** | **75-80 y** | **75-80 y** | **≥ 60 y** |
| **Coverage** | **60%** | **60%** | **60%** | **80%** | **60%** | **80%** | **60%** | **80%** |
| **Revaccination** | **No** | **No** | **Every 2 y** | **Every 2 y** | **No** | **No** | **Every 3 y** | **Every 3 y** |
| Number of vaccine doses | 8,912,674 | 10,779,577 | 39,901,688 | 152,238,080 | 19,906,760 | 11,902,883 | 27,249,055 | 106,397,904 |

**Abbreviations**: y, years.

##### *S2.5 Additional results of the exploratory analysis*

Exploratory analysis was conducted for Strategies 4 and 8 only. The observed trends in disease impact were similar for these two strategies, therefore the results for Strategy 4 are presented in the main text, and corresponding outputs for Strategy 8 are provided below (**Table S20**).

**Table S20.** Exploratory analysis: Number of cases, number of cases avoided, and percentage of cases avoided in the **target population**, mRNA-1345 vs no vaccination, over 20 years (Strategy 8)

| Strategy | No vaccination | Strategy 8 | Strategy 8 | No vaccination | Strategy 8 |
|---|---|---|---|---|---|
| Analysis | Main analysis | Main analysis | Exploratory analysis, alternative VE | Exploratory analysis, demographic shift | Exploratory analysis, demographic shift |
| **Number of cases** | | | | | |
| RSV ARD | 153,877,864 | 142,031,599 | 142,971,964 | 146,958,230 | 135,794,816 |
| RSV LRTD | 12,025,486 | 10,178,790 | 10,123,650 | 12,523,099 | 10,555,908 |
| RSV hospitalisation | 1,090,248 | 831,911 | 811,197 | 1,141,781 | 866,035 |
| RSV death | 336,077 | 191,499 | 178,969 | 358,961 | 204,501 |
| **Number of cases avoided** | | | | | |
| RSV ARD | - | 11,846,264 | 10,905,900 | - | 11,163,414 |
| RSV LRTD | - | 1,846,696 | 1,901,835 | - | 1,967,191 |
| RSV hospitalisation | - | 258,336 | 279,050 | - | 275,746 |
| RSV death | - | 144,578 | 157,108 | - | 154,461 |
| **Proportion of cases avoided** | | | | | |
| RSV ARD | - | 8% | 7% | - | 8% |
| RSV LRTD | - | 15% | 16% | - | 16% |
| RSV hospitalisation | - | 24% | 26% | - | 24% |
| RSV death | - | 43% | 47% | - | 43% |

**Abbreviations:** VE, Vaccine efficacy.

It should be noted that in the exploratory analysis on a demographic shift, a more pronounced population growth was projected in comparison to the main analysis. While in the main analysis population size increased by around 1.2 million over 20 years, in the exploratory analysis a 2.9 million increase over the same time period was simulated (see **Figure S14**). Additionally, this exploratory analysis simulated change in the age distribution, with increasing proportion of the population in the older age groups. Particularly, over 2024-2044 years, the population size in the age group of ≥60 year olds increased by 2.1 million in the main analysis, and by 2.7 million in the exploratory analysis (see **Figure S15**).

**Figure S14.** Projected population structure for main analysis and exploratory analysis in 2024, 2034, and 2044.

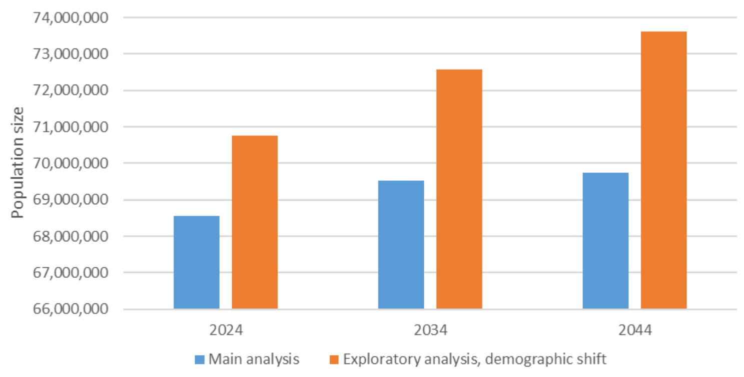


**Figure S15.** Projected population structure by age for main analysis and exploratory analysis in 2024, 2034, and 2044.

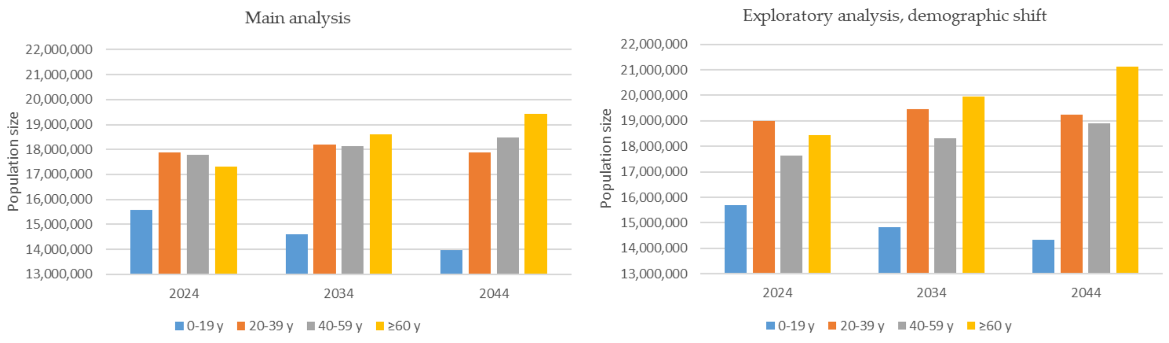


**Abbreviations**: y, years.
